# Cognitive, emotional, and social functioning of preschoolers with attention deficit hyperactivity problems

**DOI:** 10.1101/2021.02.18.21251985

**Authors:** Guido Biele, Kristin R. Overgaard, Svein Friis, Pal Zeiner, Heidi Aase

## Abstract

Attention Deficit Hyperactivity Disorder (ADHD) is associated with deficits in a number of functional domains, which have been described both as causes and effects of ADHD. It remains unclear if deficits in different domains are equally strong in early childhood, and which deficits are specific to ADHD. Here, we describe functional domains in preschoolers and assess deficits in children with ADHD problems, by comparing them to preschoolers who have other mental health problems or who develop typically. The ADHD Study assessed 1195 ca. 3.5 years old preschoolers through a semi-structured parent interview, parent questionnaires, and with neuropsychological tests. We determined functional domains by applying factor analytic methods to a broad set of questionnaire- and test-scales. Using resulting factor scores, we employed a Bayesian hierarchical regression to estimate functional deficits in children with ADHD. We found that preschoolers’ functioning could be described along the seven relatively independent dimensions activity level and regulation, executive function, cognition, language, emotion regulation, introversion, and sociability. Compared to typically developing preschoolers, those with ADHD had deficits in all domains except introversion and sociability. Only deficits in activity level and regulation and executive functions were larger than 0.5 standardized mean deviations and larger than deficits of children with other mental health problems. Preschoolers with ADHD have deficits in multiple functional domains, but only impairments in activity level and regulation and executive functions are specific for ADHD and large enough to be clinically significant. Research on functioning in these domains will likely be central for understanding the development of ADHD in later the childhood, and for improving treatment and prevention approaches.

---

Attention Deficit Hyperactivity Disorder (ADHD) is a neurodevelopmental disorder characterized by symptoms of inattention, impulsivity, and hyperactivity (Thapar and Cooper, 2016). Children with ADHD have difficulties in a number of functional domains, including cognition, language, and social behavior (Willcutt *et al*., 2008; Larson *et al*., 2011). While ADHD symptoms have been described as a cause of functional deficits (e.g. Rommel *et al*., 2015), a review of prominent ADHD theories shows that functions such as executive cognition or reward processing are also seen as causes for the development of ADHD symptoms (Ziegler *et al*., 2016). Whereas there is a general agreement that children with ADHD have difficulties in several domains, the relative strength and specificity of functional deficits in different domains remains unclear, especially in early childhood.

The current Diagnostic and Statistical Manual of Mental Disorders (DSM 5, American Psychiatric Association, 2013) prescribes assessment of functioning in the domains Cognition, Mobility, Self-care, Getting along, Life activities and Participation with the WHO Disability Assessment Schedule 2.0 (Ustun *et al*., 2010). The DSM-5 also defines five neurocognitive domains for the diagnosis of neurocognitive disorders (Sachdev *et al*., 2014), which are closer to how functional domains are described in basic research about ADHD (Nikolas and Nigg, 2013; Mueller *et al*., 2017). For this article, we define functional domains as groups of mental processes that are more closely related to each other than to mental processes from other domains.

Investigating deficits in different functional domains has been proposed to improve the understanding of mental health disorders. The Research Domain Criteria framework (RDoC, Insel *et al*., 2010) seeks to understand and explain mental health disorders as a combination of deficits across the domains negative valence systems, positive valence systems, cognitive systems, systems for social processes, and arousal/regulatory systems. Because many ADHD characteristics are thought to be extreme expressions of traits present in the general population, a dimensional approach should be especially useful for understanding ADHD (Baroni and Castellanos, 2015). While researchers have investigated neuropsychological profiles (e.g., Fair *et al*., 2012; Hulst *et al*., 2015; Lambek *et al*., 2018) or temperamental aspects of ADHD (Karalunas *et al*., 2014), there is a lack of ADHD research that examines psychological functioning across a broad range of domains. Moreover, even though early functional deficits likely play an important role in the development of ADHD symptoms, there is to date little research on functional profiles of preschoolers.

The ADHD Study, a sub-study of the Norwegian Mother and Child Birth Cohort Study (MoBa, Magnus *et al*., 2006), documented functional deficits in language, executive functions, affect and emotion regulation, and social behavior in preschoolers with ADHD (Skogan *et al*., 2014, 2015; Overgaard *et al*., 2016, 2018; Rohrer-Baumgartner *et al*., 2016; Bendiksen *et al*., 2017). While these studies demonstrate functional deficits in preschoolers with ADHD and related disorders in a number of domains, the picture remains incomplete. It is unclear if these domains are independent, if deficits compared to typically developing children are equally large in all domains, and which deficits are especially strong in ADHD compared to preschool age children with other mental health or developmental problems. Lastly, the inherent uncertainty of mental health diagnoses in preschoolers (McClellan and Speltz, 2003) raises the question if children who fulfill diagnostic criteria for symptoms *and* impairments have different functional profiles than children who only fulfill symptom criteria.

Hence, the aim of this article is to describe functional domains in preschoolers, and to explore how functional profiles of preschoolers with ADHD problems differ from profiles of children with other mental health problems and typically developing controls. We hypothesize that the broad set of measures collected in the ADHD study is organized in domains similar to those described in the RDoC approach, and that preschoolers with ADHD have the greatest impairments in domains that are related to core ADHD symptoms.

## Methods

### Study design and recruitment of participants

The Norwegian Mother and Child Cohort Study (MoBa) is an ongoing prospective population-based cohort study of Norwegian-speaking women that is conducted by the Norwegian Institute of Public Health (Magnus *et al*., 2006). The current article is from a clinical sub-study on ADHD, for which Figure S1 shows the recruitment procedure. This study oversampled children at risk for ADHD, by using data from the MoBa questionnaire that was administered to mothers at child age 3 years (Overgaard *et al*., 2014). The questionnaire included 11 items about ADHD, including six items from the Child Behavior Checklist/1.5-5 (CBCL, Achenbach and Rescorla, 2000) and five items from the DSM-IV-TR criteria for ADHD (American Psychiatric Association, 2000). Children were identified as at risk for ADHD when their sum score from the 11 ADHD symptom questions was at or above the 90th percentile of scores from children in MoBa who were born before 2004, or when the mother indicated hyperactivity-related health problems. In addition to 2801 preschoolers at risk for ADHD, 651 control participants were invited to participate. In total, about 35% agreed to participate in the ADHD Study, and from 2007 to 2011, 1195 children (mean age: 3.5 years, age range: 3.1 to 3.8 years) took part in a 1-day clinical assessment.

## Material

The study used the semi-structured clinical parent-interview “Preschool Age Psychiatric Assessment” (PAPA, Egger and Angold, 2004) to assess mental health symptoms. The PAPA interview elicits, based on DSM IV diagnostic classifications, information about symptoms and impairments for many mental health disorders of the childhood. This includes ADHD, conduct disorder, oppositional defiant disorder, and anxiety disorders. In the ADHD study, only symptoms lasting longer than 3 months were counted as present. The ADHD Study modified the PAPA by adding a section about impairments for each mental health problem (details in the supplementary materials). As an inter-rater reliability check, a separate rater who was blind to the parent and teacher screen ratings re-scored audiotapes of 79 randomly selected assessment interviews. The average intra-class correlations (ICCs) were .97 for HI symptoms, .99 for IA symptoms, and .98 for the total number of ADHD symptoms.

This research used data from following parent questionnaires: The Behavior Rating Inventory of Executive Function–preschool version (BRIEF–P, Gioia *et al*., 2002, 2007) assesses functional deficits on the sub-scales *inhibit, shift, emotional control, working memory*, and *plan and organize*. From the language section of the Child Development Inventory (CDI, Ireton and Glascoe, 1992, 1995) we used 50 items to score expressive language. The Children’s Behavior Questionnaire (CBQ, Putnam and Rothbart, 2006) is a temperament questionnaire with 50 items grouped into the sub scales *activity level, anger, attention focusing, discomfort, fear, high intensity pleasure, low intensity pleasure, impulsivity, inhibitory control, perceptual sensitivity, sadness*, and *soothability*. The Emotionality, Activity and Shyness temperament questionnaire (EAS, Buss and Plomin, 1984; Mathiesen and Tambs, 1999) uses 12 items to score preschoolers on the sub-scales *activity, emotion, shyness*, and *sociability*.

The ADHD Study used a sub-set of the NEuroPSYchological Assessment Battery tests (NEPSY, Korkman *et al*., 1998, 2000), including those to assess *inhibition, visual attention, visuospatial processing*, and *language*. The Norwegian version of the Boston Naming Test (BNT, Kaplan *et al*., 1983) assess expressive language. From the Stanford-Binet Intelligence Scales 5th ed. (Roid, 2003) the ADHD study used the Verbal Memory for Sentences test (VMS) as an indicator of *verbal working memory*, the Delayed Response task/Block Span test (DR/BS) as an indicator of *nonverbal working memory*, the Object series, Pattern analysis/Matrices tests (OS/PAM) as an indicator of *nonverbal IQ* and the Comprehension/Vocabulary tests (CM) as an indicator for *verbal IQ*. Finally, the ADHD study used a version of the Cookie Delay Task (CDT) to assess delay aversion. See the supplementary information for further details about the instruments.

### Procedure

Parents who had consented to participate received questionnaires around 4 weeks before the assessment day. For the clinical assessment, all participants traveled to Oslo University Hospital. At the assessment day clinical psychologists or psychiatrist conducted neuropsychological tests and medical examinations in dedicated rooms. Trained graduate students in psychology conducted the PAPA parent interview under supervision of clinical experts. The assessment day concluded with a debriefing for parents.

The present study was approved by the Norwegian Regional Ethics Committee for Medical and Health related Research.

### Data analysis

All analyses were performed with R (R Core Team, 2019) or Mplus (Muthen and Muthen, 2017). The supplementary information contains additional analysis details.

### Classification of participants into problem groups

Classifications were determined by an algorithm that applied DSM IV symptom and impairment criteria to PAPA data. Children were classified as having ADHD, behavioral (Oppositional Defiant or Conduct Disorder) or anxiety (Social, Separation, or Generalized Anxiety, Phobia) problems with impairment, if they fulfilled DSM IV symptom and impairment criteria. They were classified as having problems without impairment, if they fulfilled only symptom criteria. We also used clinicians’ global evaluation of the presence and impairing nature of language problems. Preschoolers who had multiple but only one impairing problem, were classified based on this problem. Children with multiple impairing problems were classified into the first of the following problem-groups for which they fulfilled diagnostic criteria: 1. ADHD, 2. behavior, 3. anxiety, and 4. language problems. Children without a mental health problem were categorized as typically developing. (See also Figures S3 and S4.)

### Choice and scoring of tests and scales

We included scales and tests that measure psychological functioning and for which data from at least 70% of the children was available. These are the abbreviated sub-scales of the Stanford Binet test, NEPSY, CDT, BNT, the language section of the CDI, and all sub-scales from the BRIEF-P, CBQ, EAS questionnaires.

All tests except the BNT were scored based on the published test manuals. BNT and questionnaire sub-scales were scored by estimating Rasch Item Response Theory models (Rasch, 1960) for each sub-scale of a questionnaire.

### Identification of functional domains through Exploratory Structural Equation Modeling

This step used test- and trait-scores calculated in the previous step. An initial analysis used a confirmatory factor analysis to examine if a theory-based assignment of scales to RDoC domains fitted the data well. For the data driven identification of functional domains we estimated exploratory structural equation models (ESEM). Lastly, we used CFA to explore if the model structure of the best ESEM factor model could be simplified by setting weak cross loadings to zero. Factor scores from the final model capture individual functioning in the different domains and were basis for further analyses.

### Comparison of functioning across functional domains and groups

We employed a Bayesian hierarchical linear regression model to examine associations of mental health problems and functioning in all domains simultaneously, while also taking into account that associations might vary across gender and groups with different problems and degrees of impairment. By estimating the model in a Bayesian framework with weakly informative priors, one can reliably estimate variation in effect sizes between groups, and automatically control the multiple comparison problem (Gelman *et al*., 2012).

Consistent with recent recommendations about statistical practice (Sullivan and Feinn, 2012; Wasserstein *et al*., 2019) we report effect size means and the 90% credible intervals instead of p-values. To communicate the probability of clinically significant impairments or differences between groups, we report posterior probabilities that a difference is larger than 0.5 standardized mean differences (SMD).

### The study sample

Parents of 40% of at risk for ADHD and 25% of control children consented to participate. Altogether 1195 children participated in the assessment, of which 1184 children had less than 50% of missing data on tests and scales of interest. Of these participants, 219 fulfilled DSM IV ADHD symptom criteria (168 with and 51 without impairment). Table 1 describes the study sample.

**Table 1.**
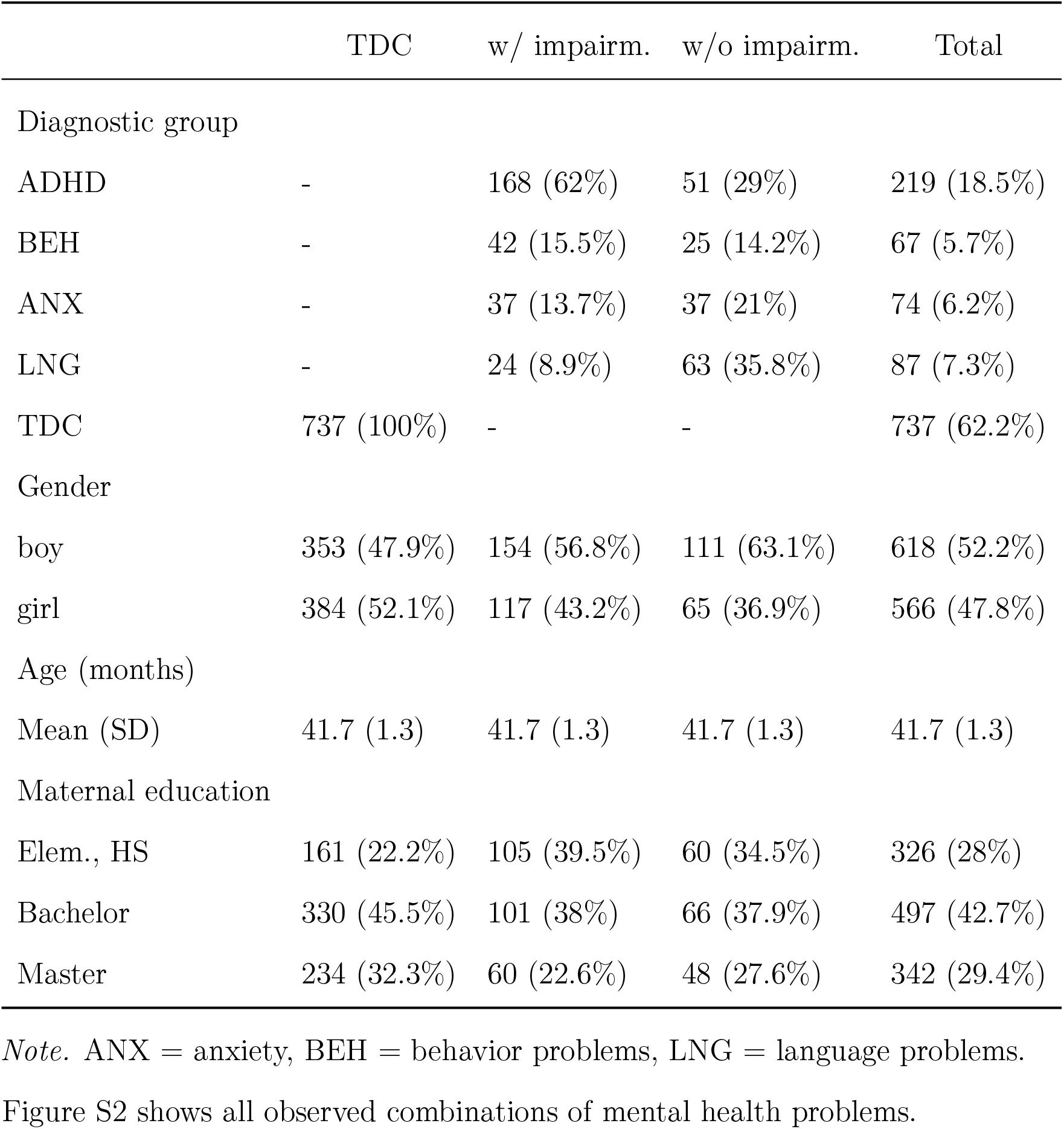
Study sample

|  | TDC | w/ impairm. | w/o impairm. | Total |
| --- | --- | --- | --- | --- |
| Diagnostic group |  |  |  |  |
| ADHD | - | 168 (62%) | 51 (29%) | 219 (18.5%) |
| BEH | - | 42 (15.5%) | 25 (14.2%) | 67 (5.7%) |
| ANX | - | 37 (13.7%) | 37 (21%) | 74 (6.2%) |
| LNG | - | 24 (8.9%) | 63 (35.8%) | 87 (7.3%) |
| TDC | 737 (100%) | - | - | 737 (62.2%) |
| Gender |  |  |  |  |
| boy | 353 (47.9%) | 154 (56.8%) | 111 (63.1%) | 618 (52.2%) |
| girl | 384 (52.1%) | 117 (43.2%) | 65 (36.9%) | 566 (47.8%) |
| Age (months) |  |  |  |  |
| Mean (SD) | 41.7 (1.3) | 41.7 (1.3) | 41.7 (1.3) | 41.7 (1.3) |
| Maternal education |  |  |  |  |
| Elem., HS | 161 (22.2%) | 105 (39.5%) | 60 (34.5%) | 326 (28%) |
| Bachelor | 330 (45.5%) | 101 (38%) | 66 (37.9%) | 497 (42.7%) |
| Master | 234 (32.3%) | 60 (22.6%) | 48 (27.6%) | 342 (29.4%) |

*Note.* ANX = anxiety, BEH = behavior problems, LNG = language problems.

## Results

### Dimensions of functioning

Initial CFAs, for which sub-scales were on theoretical grounds assigned to RDoC domains, did not describe the data sufficiently well (see Table S1). The final ESEM analysis showed that a 7-factor model was the simplest model that could adequately describe the data (RMSEA = 0.03 (0.028, 0.037), CFI = 0.96). Because follow up CFA analyses that constrained small cross-loadings to 0 resulted in unsatisfactory RMSEA and CFI statistics, we retained the final ESEM model as the best model of functional domains.

Table S2 and Figure S5 show the factor loadings from the best ESEM model. This suggests following factors: *activity level and regulation* (AL, the ability to down-regulate physical activity), *executive functions* (EF, the ability to regulate behavior and cognitive functions), *cognition* (CO, working memory and cognitive flexibility), *language* (LA), *emotion regulation* (ER, the ability to control emotions), *introversion* (IN, the preference for calm activities), and *sociability* (SO, the ability to get into contact with others).

The average of the unsigned factor correlations was 0.15. Following factor correlations were larger than 0.30: r(LA,CO) = 0.39, r(EF,AL) = 0.32, r(ER,EF) = 0.32 (c.f. Table S3).

### Functional profiles of preschoolers with ADHD problems

Across all domains, functioning of preschoolers with ADHD was −0.42 standardized mean differences (SMD) below the functioning of typically developing preschoolers. The average SMDs for preschoolers with ADHD with and without impairment were −0.55 and −0.30, respectively. Preschoolers with language problems had functioning deficits similar to preschoolers with ADHD, whereas children with behavior or anxiety problems had milder deficits (c.f. Table S4).

Figure 1 and Table 2 show functional profiles. Preschoolers with ADHD were particularly impaired in the domains EF and AL, where their functioning was around −0.90 SMD below that of typically developing controls (*P* (*SMD < −*0.5)=1). In comparison, functioning levels of preschoolers with ADHD in the domains CO, LA and ER was only around −0.50 SMD below typically developing controls (*P* (*SMD < −*0.5)=0.56).

**Figure 1.**
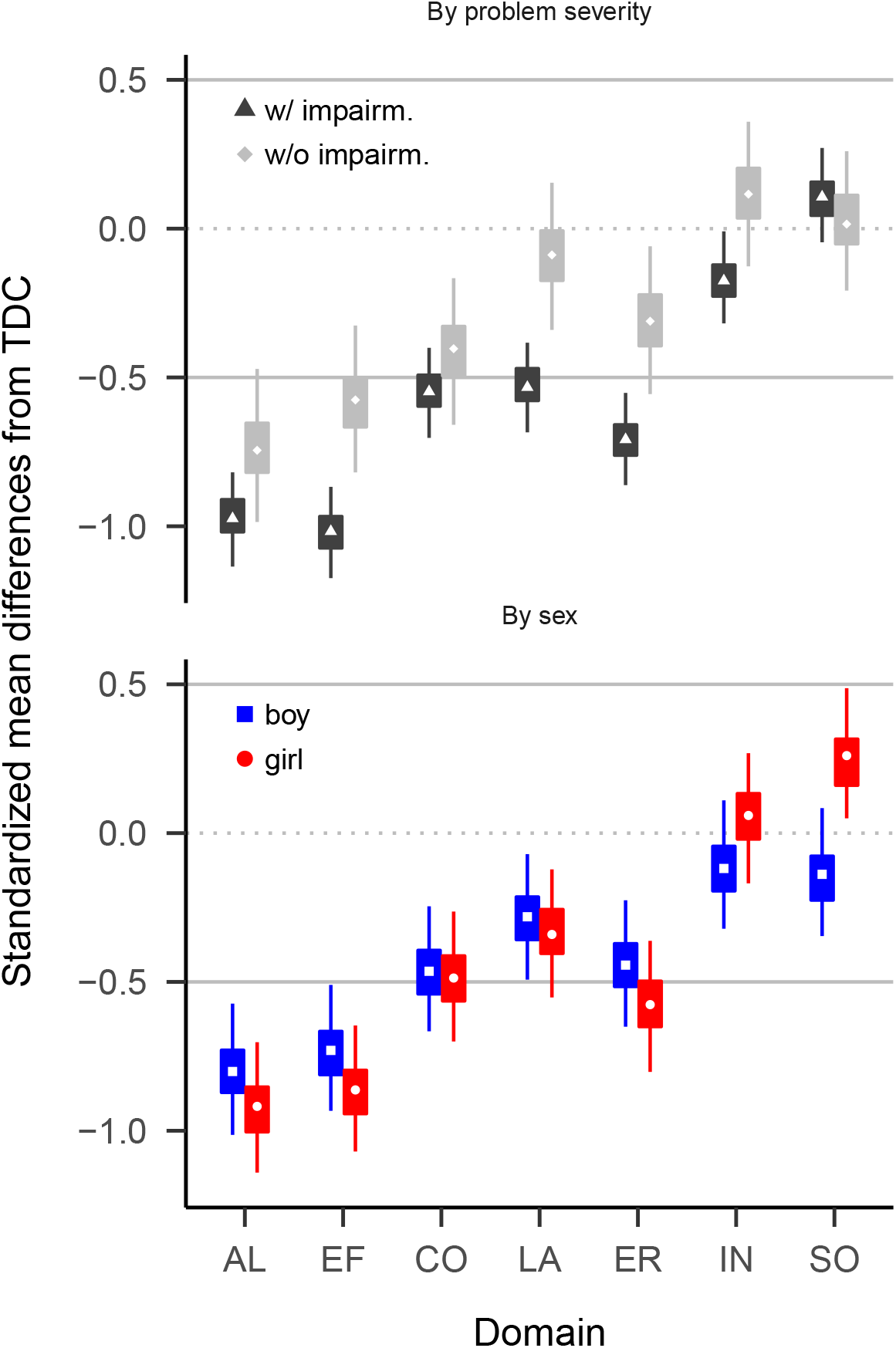
Comparison of preschoolers with ADHD with typically developing preschoolers (TDC). Values below zero indicate deficits in preschoolers with ADHD. Boxes cover 50% highest density intervals (HDIs), thin vertical lines cover 90% HDIs. The two solid horizontal lines enclose a range from −0.5 to 0.5 SMD.

**Table 2.** Differences in functioning between preschoolers with ADHD and preschoolers with no or other problems.

|  | TDC vs |  |  | other MHP vs |
| --- | --- | --- | --- | --- |
|  | ADHD w/ impairm. | ADHD w/o impairm. | All ADHD | All ADHD |
| AL | -0.97 (-1.14, -0.82; 1.00) | -0.74 (-0.99, -0.47; 0.97) | -0.92 (-1.07, -0.78; 1.00) | -0.89 (-1.41, -0.37; 0.88) |
| EF | -1.02 (-1.17, -0.87; 1.00) | -0.58 (-0.82, -0.33; 0.74) | -0.91 (-1.04, -0.77; 1.00) | -0.48 (-0.92, -0.07; 0.44) |
| CO | -0.55 (-0.70, -0.40; 0.73) | -0.40 (-0.66, -0.17; 0.22) | -0.51 (-0.65, -0.37; 0.58) | -0.28 (-0.81, 0.32; 0.28) |
| LA | -0.53 (-0.68, -0.38; 0.67) | -0.09 (-0.34, 0.15; 0.00) | -0.43 (-0.55, -0.28; 0.16) | 0.36 (-0.41, 1.31; 0.02) |
| ER | -0.71 (-0.86, -0.55; 0.99) | -0.31 (-0.56, -0.06; 0.07) | -0.62 (-0.75, -0.47; 0.95) | -0.12 (-0.62, 0.33; 0.1) |
| IN | -0.18 (-0.32, -0.01; 0.00) | 0.12 (-0.13, 0.36; 0.00) | -0.11 (-0.24, 0.04; 0.00) | 0.14 (-0.34, 0.58; 0.01) |
| SO | 0.11 (-0.05, 0.27; 0.00) | 0.02 (-0.21, 0.26; 0.00) | 0.09 (-0.07, 0.22; 0.00) | 0.47 (-0.06, 1.21; 0) |

*Note.* Standardized mean deviations (SMD). Values are mean (lower, upper 90% HDI; P(SMD<-0.5)). See main text for abbreviations LA - SO.

Functioning in the domains SO and IN did not differ substantially from typically developing controls (average SMD=0; *P* (*SMD < −*0.5)=0). Preschoolers with ADHD with and without impairment had similar functioning patterns. However, with exception of the domains CO and SO, those with impairment had around 0.30 SMD larger deficits than those without. We found overall small gender differences. Only in SO did boys with ADHD show somewhat weaker deficits than girls.

Figure 2 and Tables 2 and S7 report comparisons between preschoolers with ADHD and those with other mental health or developmental problems. Only deficits in AL (with impairment: SMD = −1.04 (−1.23, −0.86; P(SMD<-.5)=1.00), without impairment: SMD = −0.74 (−0.98, −0.49; P(SMD<-.5)=0.94)) and EF (with impairment: SMD = −0.61 (−0.79, −0.42; P(SMD<-.5)=0.84)), without impairment: SMD = −0.35 (−0.59, −0.11; P(SMD<-.5)=0.15)) were larger in preschoolers with ADHD, compared to preschoolers with other mental health problems. Preschoolers with ADHD had only moderately more difficulties in CO (with impairment: SMD = −1.04 (−1.23, −0.86; P(SMD<-.5)=1.00), without impairment: SMD = −0.74 (−0.98, −0.49; P(SMD<-.5)=0.94)) compared to preschoolers with other mental health problems, and similar or fewer difficulties in the domains LA, ER, IN, or SO (with impairment: SMD = 0.14 (0.04, 0.25; P(SMD<-.5)=0.00)), without impairment: SMD = 0.28 (0.15, 0.41; P(SMD<-.5)=0.00)). In particular, children with ADHD had much weaker LA deficits than preschoolers with impairing language problems (with impairment: SMD = 1.20 (0.89, 1.53; P(SMD<-.5)=0.00), without impairment: SMD = 0.86 (0.58, 1.14; P(SMD<-.5)=0.00)), and were more sociable than children with impairing anxiety problems (with impairment: SMD = 1.12 (0.87, 1.38; P(SMD<-.5)=0.00), without impairment: SMD = 0.82 (0.52, 1.13; P(SMD<-.5)=0.00)). Figure 3 summarises the comparisons and highlights that, whereas preschoolers with ADHD are impaired compared to TDC in most functional domains, only impairments of AL and EF are specific to ADHD in that they are larger than 0.5 SMD compared to TDC and also larger than deficits of preschoolers with other mental health problems. See Table S6 for more details.

**Figure 2.**
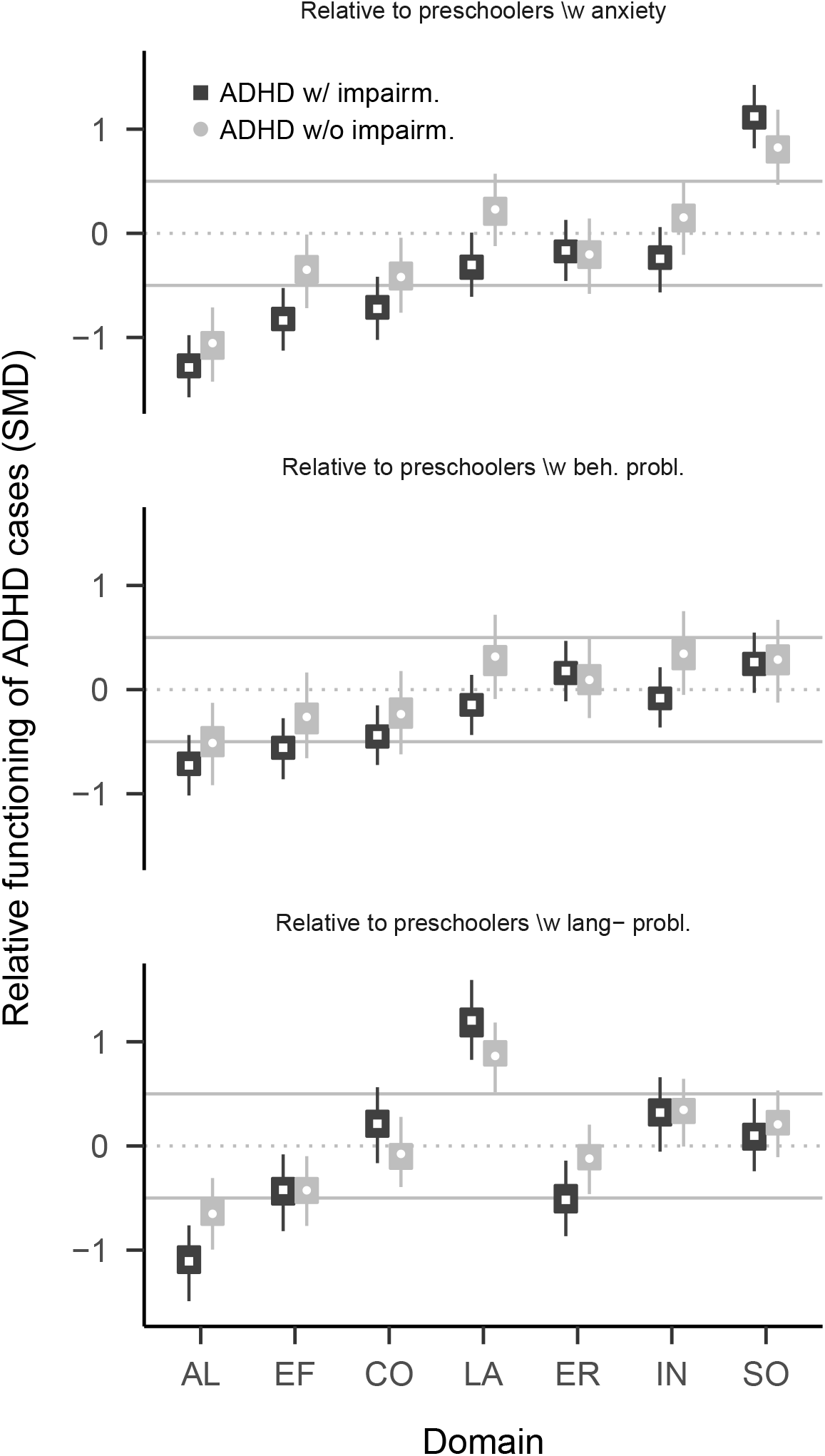
Comparison of preschoolers with ADHD with preschoolers with other mental health problems. Relative functioning is calculated as the factor score difference between ADHD group and comparison group. Values below zero indicate that the comparison group has less problems than preschoolers with ADHD.

**Figure 3.**
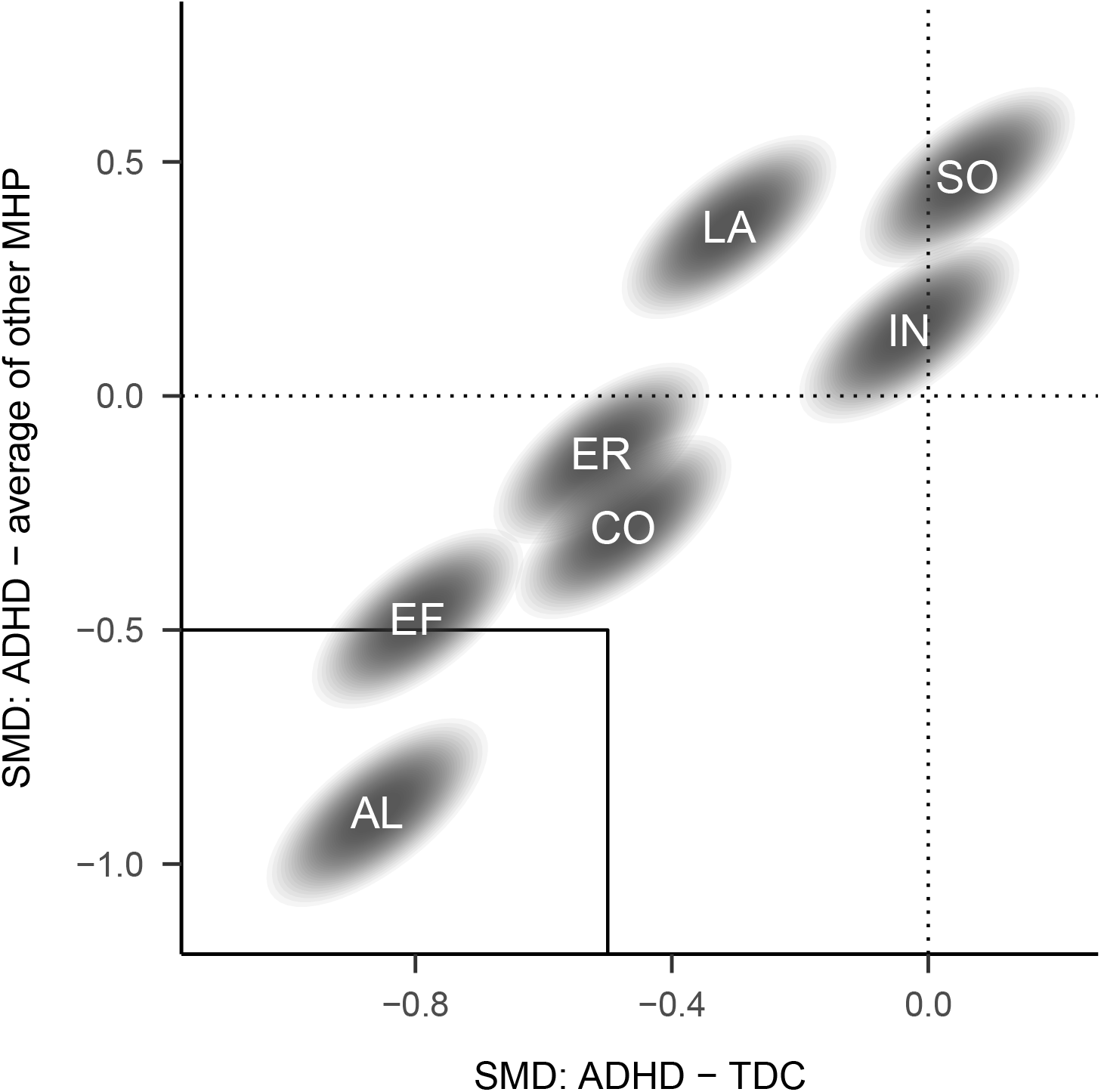
Joint comparison of factor scores for preschoolers with ADHD with typically developing preschoolers (TDC, on the x-axis) and those with other mental health problems (MHP, on the y-axis). Differences are averaged across preschoolers with and without impairment. Values below zero indicate that preschoolers with ADHD have stronger problems than the comparison group. Ellipses cover 90% highest density intervals. Labels in ellipses indicate functional domains. Domains in the lower left rectangle are those where preschoolers with ADHD have a 0.5 SMD large deficit compared to TDC and children with other MHPs. See Figure S10 for comparisons with specific other MHPs.

## Discussion

The aim of the current study was to describe domains of functioning in preschoolers and to compare functioning of preschoolers with ADHD problems with that of preschoolers with other or no problems. We identified seven weakly correlated functional domains: Executive functioning (EF), activity level and regulation (AL), cognition (CO), language (LA), introversion (IN), emotion regulation (ER) and sociability (SO). Compared to typically developing children, functional deficits of preschoolers with ADHD were especially pronounced in AL and EF, less pronounced in CO, LA, and ER, and absent in SO and IN. Compared to children with anxiety, behavior or language problems, children with ADHD had similar or weaker deficits in all domains except AL and EF, where they had substantially greater impairments. Therefore, only EF and AL impairments appear specific to children with ADHD problems.

To identify functional domains, we used results from a broad range of instruments, including neuropsychological and performance tasks, and parent-questionnaires about cognitive functioning and temperament (personality traits). We chose this range of measures, because both performance-based and temperamental traits can contribute to mental health problems (Fair *et al*., 2012; Karalunas *et al*., 2014). More generally, temperamental and performance-based traits of individuals are relatively stable over time and correlated with each other (Ackerman and Heggestad, 1997). Therefore, we reason that joint examination of temperament and performance based aspects of behavior are important for understanding ADHD and its development. This view is also supported by our finding that that preschoolers with ADHD have deficits in the temperamental (Activity level and regulation) and the performance domain (executive functions).

A likely reason for divergences between our and the RDoC domains is that the set of instruments used influences the what domains a study recovers. The use of many instruments to assess language and cognition likely contributed to our identification of two domains in this area, whereas the sparse assessment of reward processing likely impeded us from identifying such a domain or even specific concepts within reward processing like reinforcement learning of delay aversion. Still, we view the domains identified in the ADHD Study data as broadly consistent with the RDoC domains.

Similar to earlier investigation, we observed only small correlations between assessments of the same construct with parent-questionnaires and tests. Toplak *et al*. (2013) hypothesized that the low correlation could be explained by the fact that tests primarily measure the ability to perform a task, whereas questionnaires in addition measure the ability to independently focus energy in a goal-oriented manner. This interpretation assumes that both instrument types reliably measure traits and abilities. The low correlations could also be due to unsystematic and systematic errors in both instrument types. For example, tests can measure state in addition to trait, and questionnaires suffer from response biases or varying understanding fo parents about what behavior is normative. Repeated administration of tests would provide the most reliable information and should if possible be used in the assessment of children.

As expected, we found the largest deficits of children with ADHD in the domains of activity level and regulation and executive functions. The prominence of EF and AL impairments is consistent with a dual pathways view of the development of ADHD, which suggests that ADHD can be caused by earlier functional deficits (Sonuga-Barke, 2003). While the original formulation of the dual pathway hypothesis highlights deficits in EF and reward processing as key causes for the development of ADHD symptoms, we only find direct evidence for early EF deficits, likely due to the sparse assessment of reward processing in the ADHD study. Still, our results provide indirect evidence for the early reward processing deficits, if one considers that some ADHD theories suggest that a heightened activity level is an indicator for impaired reward processing (Sagvolden *et al*., 2005; Ziegler *et al*., 2016). Still, future studies should put more emphasis on the assessment of reward processing in early childhood, in order to improve the understanding of its role on the development of ADHD symptoms. The prominence of AL and EF deficits in early childhood raises the likelihood that they are causes of later deficits in the other domains.

While the broadness of functions investigated in the ADHD Study and the large sample size set this study apart, some aspects of the study suggest caution when generalizing results to the broader population. The sample composition, which is characterized by self-selection into the study based on high parental education, high parental ratings of preschool ADHD symptoms, and absence of children with ASD (which were recruited into a sister study on ASD, (Stoltenberg *et al*., 2010)), does not fully reflect the population of preschoolers with ADHD problems. Regarding the absences of children with ASD, additional comparisons based on the ABC study sample shown in the supplementary materials (Figures S11 and following) suggest that the differences between children with and without ADHD we found are not unique to the ADHD Study sample. Still, given the sample characteristics, the presented results generalize most readily to a population of preschoolers at risk to develop ADHD and with relatively well-educated parents.

The current study is of exploratory nature and reports only cross-sectional associations between different functional domains. Nevertheless, the presented results suggest that because impairments in AL and EF are much stronger and central than other deficits, first choice treatment of preschoolers with ADHD from populations similar to the study sample should focus on these areas, because other functional deficits will typically be less severe. Future research on cross-lagged associations between functioning in different domains (c.f. Arnett *et al*., 2012) is needed to investigate the causal role of EF and AL for later development, and to inform if focus of treatment in these domains is indicated.

In sum, the current study identified functional domains similar to, but not identical with the RDoC framework. Preschoolers with ADHD have deficits in most functional domains, but only deficits in activity level and regulation and executive functions were clinically significant (SMD > 0.5) and more expressed in preschoolers with ADHD compared to those with other problems. Future longitudinal research on the development of functioning in domains over time will be important to investigate a causal role of early functional deficits in the development of ADHD, and for the further development of effective early interventions for ADHD.

## Supporting information

Supplementary methods and results

## Data Availability

Data can be obtain through application to the Norwegian Mother, Father and Child Cohort Study (MoBa)
https://www.fhi.no/en/studies/moba/

