## Supplementary methods and results for "Cognitive, emotional, and social functioning of preschoolers with attention deficit hyperactivity problems"

Supplementary Material A

### Recruitment

Figure S1 desribes the recruitment and inclusion procedure.

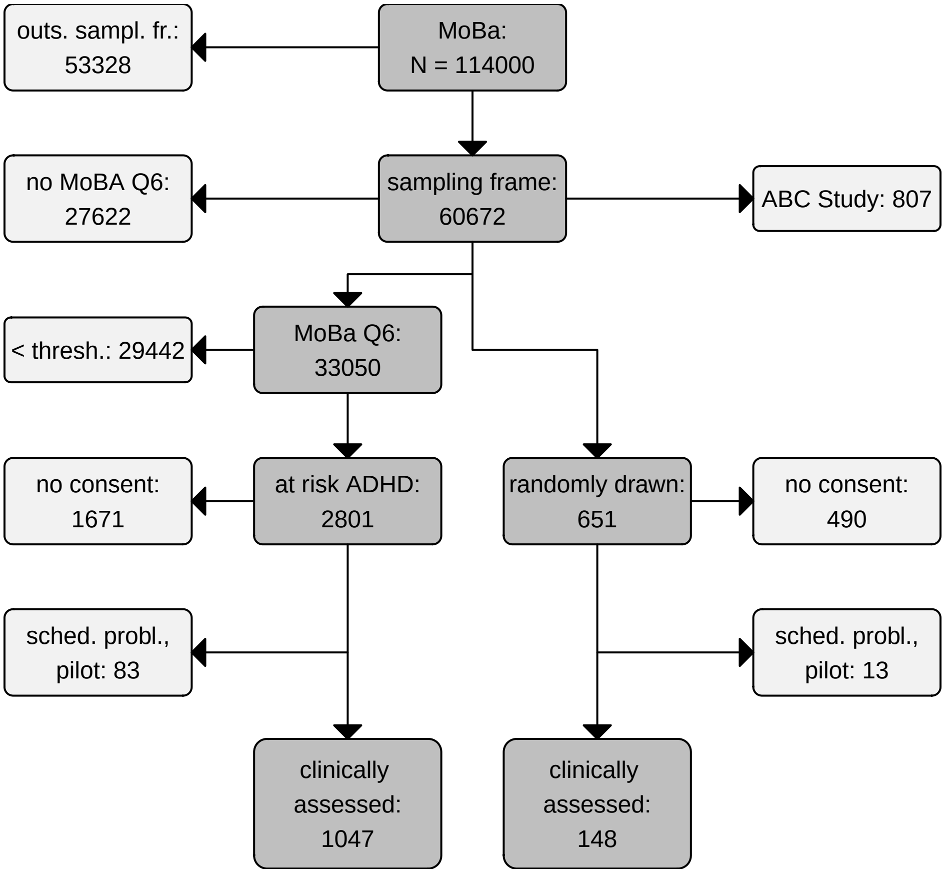


*Figure S1.* Recruitment flow chart. The inclusion period for the ADHD study lasted from April 2004 to January 2008. Q6 is the MoBa questionnaire sent out at child age three years. Participants of the ABC Study (Stoltenberg *et al.*, 2010) were not eligible for the ADHD Study. “thresh.” is the cut-off sum-score for identification of preschoolers at risk for ADHD. Some participants who consented participated in pilot assessments, and for some there was no assessment due to scheduling problems.

**Full list of instruments**

The ADHD study used the semi-structured clinical interview “Preschool Age Psychiatric Assessment” [PAPA; Egger *et al.* (2006)] to assess mental health symptoms. The PAPA interview elicits, based on DSM IV diagnostic classifications, information about the severity and frequency of symptoms and impairment for many mental health disorders of the childhood including ADHD, conduct disorder, oppositional defiant disorder, and anxiety disorders.

Parents and kindergarten teachers filled out the following questionnaires:

- Strength and Difficulties Questionnaire (SDQ, used from 2007-2009, Goodman, 1999)
- Conners’ Rating Scale – Revised, Short version, for parents (CPRS-RS, used from 2007-2009, Conners *et al.*, 1998)
- Early Childhood Inventory 4 [ECI4, used from 2010-2012; Sprafkin *et al.* (2002)]
- Behavior Rating Inventory of Executive Function – preschool version (BRIEF-P, Gioia *et al.*, 2002, 2007)
- and the language section of the Child Development Inventory (CDI, Ireton and Glascoe, 1992, 1995).

In addition, only parents received the

- Children’s Behavior Questionnaire (CBQ, Putnam and Rothbart, 2006)
- Emotionality, Activity and Shyness temperament questionnaire (EAS, Buss and Plomin, 1984; Mathiesen and Tambs, 1999)
- and also responded again to the 11 screening questions used to detect ADHD cases.

Only teachers received the Preschool Play Behavior Scale (PPBS, Coplan and Rubin, 2001).

To facilitate cooperation with a sister-study, some questionnaire instruments were exchanged half way through the study. Until 2009 parents and teachers received the SDQ and the CPRS-RS and from 2010 on the ECI-4 and some impairment questions from the SDQ. Around the same time, the wording of few BRIEF-P items were updated, which had however no effect on the scale performance (Skogan *et al.*, 2016).

As neuropsychological tests the ADHD study employed the

- Developmental NEuroPSYchological Assessment Battery (Korkman *et al.*, 1998, 2000),
- Spin The Pots and Truck Reversal Learning tests (Hughes and Ensor, 2005)
- Grooved Pegboard Task from the Wisconsin motor steadiness battery (Matthews and H., 1978),
- Cookie Delay Task,
- Norwegian version of the Boston Naming test (BNT, Kaplan *et al.*, 1983).
- From the Stanford-Binet Intelligence Scales 5th ed. (Roid, 2003) the ADHD study used the Verbal Memory for Sentences test (VMS) to assess verbal working memory (VWM), the Object series, Pattern analysis/Object matrices tests (OS/PAM) to assess nonverbal IQ (NVIQ), the Comprehension/Vocabulary tests (CM) to assess verbal IQ (VIQ), and a task for verbal task fluid reasoning (VOS).

**Description of Cookie Delay Task.** The Cookie delay task (Campbell *et al.*, 1982) involved placing a cookie under one of several upturned transparent cups. The child was instructed to wait for a signal (clap) before retrieving the treat. Eight trials were given in a pseudo-random order with delays of between 5 and 30 seconds. The experimenter's hands were raised at the midpoint of the delay period ready to clap. The child was given one practice trial. The scoring was 0 - not inhibited, 1- partially inhibited, and 2 - fully inhibited, the total range was 0-16.

### Supplementary methods

All analysis were performed with R [Version 4.0.2; R Core Team (2017)][1] or Mplus (Muthen and Muthen, 2017). Scripts for all analysis steps are available at https://github.com/gbiele/compfunc.

#### Classification of mental health problems.

##### *Criteria for diagnostic classification.*

Mental health problems were classified according to DSM IV symptom criteria. In particular, criteria are

- for ADHD: At least 6 symptoms in either the inattention or hyperactivity-impulsivity domain

- for behavior problems: (a) Conduct Disorder: At least three symptoms, (b) Oppositional defiant disorder: At least four symptoms.

[1] We, furthermore, used the R-packages *arsenal* [Version 3.5.0; Heinzen *et al.* (2018)], *brms* [Version 2.14.4; Bürkner (2017a); Bürkner (in press)], *car* [Version 3.0.10; Fox and Weisberg (2019); Fox *et al.* (2018)], *carData* [Version 3.0.4; Fox *et al.* (2018)], *data.table* [Version 1.13.6; Dowle and Srinivasan (2018)], *diagram* [Version 1.6.5; Soetaert (2017)], *flextable* [Version 0.6.2; Gohel (2019a)], *ggplot2* [Version 3.3.3; Wickham (2016)], *Gmisc* [Version 1.11.0; Gordon (2018)], *haven* [Version 2.3.1; Wickham and Miller (2018)], *HDInterval* [Version 0.2.2; Meredith and Kruschke (2018)], *htmlTable* [Version 2.1.0; Gordon *et al.* (2018)], *knitr* [Version 1.31; Xie (2015)], *lattice* [Version 0.20.41; Sarkar (2008)], *mice* [Version 3.13.0; van Buuren and Groothuis-Oudshoorn (2011)], *MplusAutomation* [Version 0.8; Hallquist and Wiley (2018)], *officer* [Version 0.3.16; Gohel (2019b)], *papaja* [Version 0.1.0.9997; Aust and Barth (2018)], *plotrix* [Version 3.8.1; J (2006)], *psych* [Version 2.0.12; Revelle (2018)], *RColorBrewer* [Version 1.1.2; Neuwirth (2014)], *Rcpp* [Version 1.0.6; Eddelbuettel and François (2011); Eddelbuettel and Balamuta (2017)], *reshape* [Version 0.8.8; Wickham (2007)], *rstanarm* [Version 2.21.1; Stan Development Team (2016a)], *shape* [Version 1.4.5; Wickham (2007); Soetaert (2018)], *sjPlot* [Version 2.8.7; Lüdecke (2019)], *StanHeaders* [Version 2.21.0.7; Stan Development Team (2016b)], *stringi* [Version 1.5.3; Gagolewski (2018)], *stringr* [Version 1.4.0; Wickham (2018)], and *tableone* [Version 0.12.0; Yoshida and Bohn. (2018)].

- for anxiety problems: (a) Specific phobia: At least one symptom, (b) social phobia: At least one symptom, (c) separation anxiety: At lest three symptoms (d) “Generalized Anxiety, worries” A strong worries combined with physical symptoms

We further distinguished children with and without impairments, based on a section about impairments the ADHD Study had added to each Disorder section of the PAPA manual.

*Assessment of impairment.*

For each mental health problem, impairments were assessed with questions about the impact of the symptoms on:

- the child’s ability to get along with the parents an the rest of the family
- the child’s ability to keep friends
- the child’s ability to learn or work in kindergarten
- the child’s ability to participate in play or other activities outside the kindergarten
- the child’s quality of life
- the family (is the child a burden for the parents or the rest of the family)

Each impairment was assessed on a scale from 0-3: “not at all”" (score 0), “a bit” (1), “a good deal” (2) , “a lot” (3).

Symptoms were classified as impairing if at least 2 questions were scored with 1, or if at least 1 questions was scored with 2 or higher.

The PAPA interview does not have a section for language impairments. However, in the ADHD Study clinicians evaluated language problems (Expressive language problems, combined expressive and phonological problems, phonological problems) as clinical (with impairment) or sub-clinical (without impairment).

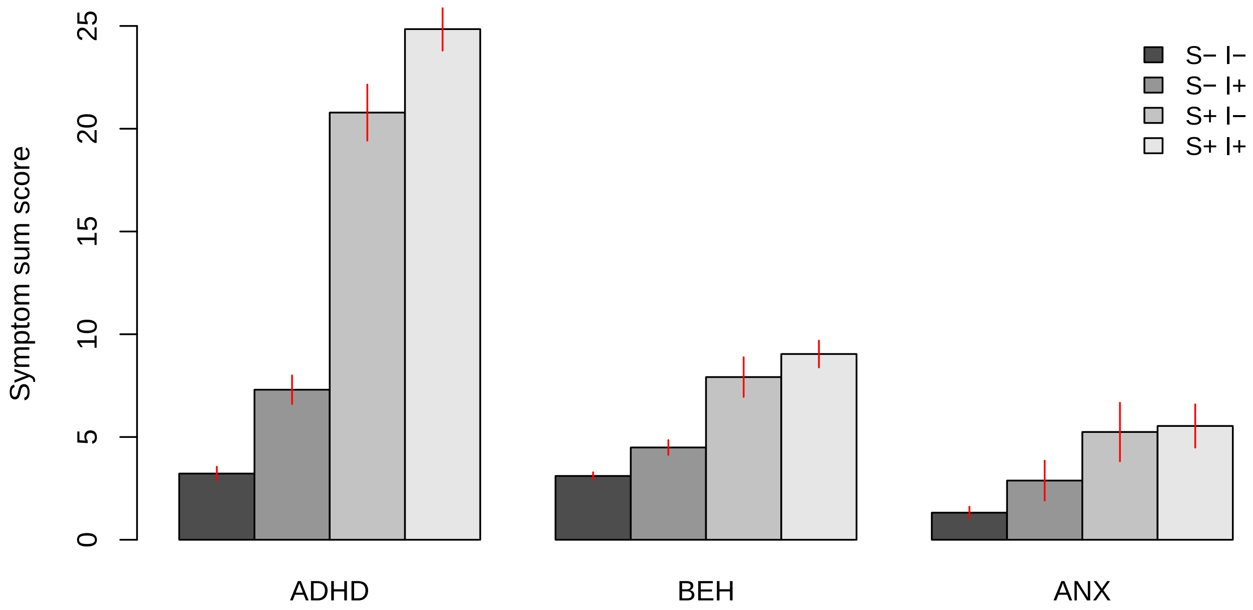


*Figure S2.* Symptoms sum scores for different diagnostic groups. "S− I−" = below symptom threshold, no impairment,"S− I+" = below symptom threshold, with impairment ,"S+ I−" = above symptom threshold, no impairment ,"S+ I+" = above symptom threshold, with impairment. As children are more clearly grouped by symptom load than by impingement, it is reasonable that symptom criteria play a greater role for classification. ADHD = Attention Deficit Hyperactivity Disorder, BEH = Behaviour problems, ANX = Anxiety problems.

**Calculation of scores for instrument sub-scales.** Scores for the NEPSY and Stanford Binet were calculated following standard procedures described in the manuals.

Scores for sub-scales of questionnaires were obtained by estimating latent Rasch models (Rasch, 1960) with the edstan packages (Furr, 2017). We used this approach as opposed to simply summing up scores and transforming them based on a norm-table, because Rasch models take better into account differences in item-difficulty and -informativeness. Moreover, norm-samples for many of the employed test are not from Europe or Norway, making the validity of normed scores from these tests for the ADHD Study questionable. Further, the explicit assumptions

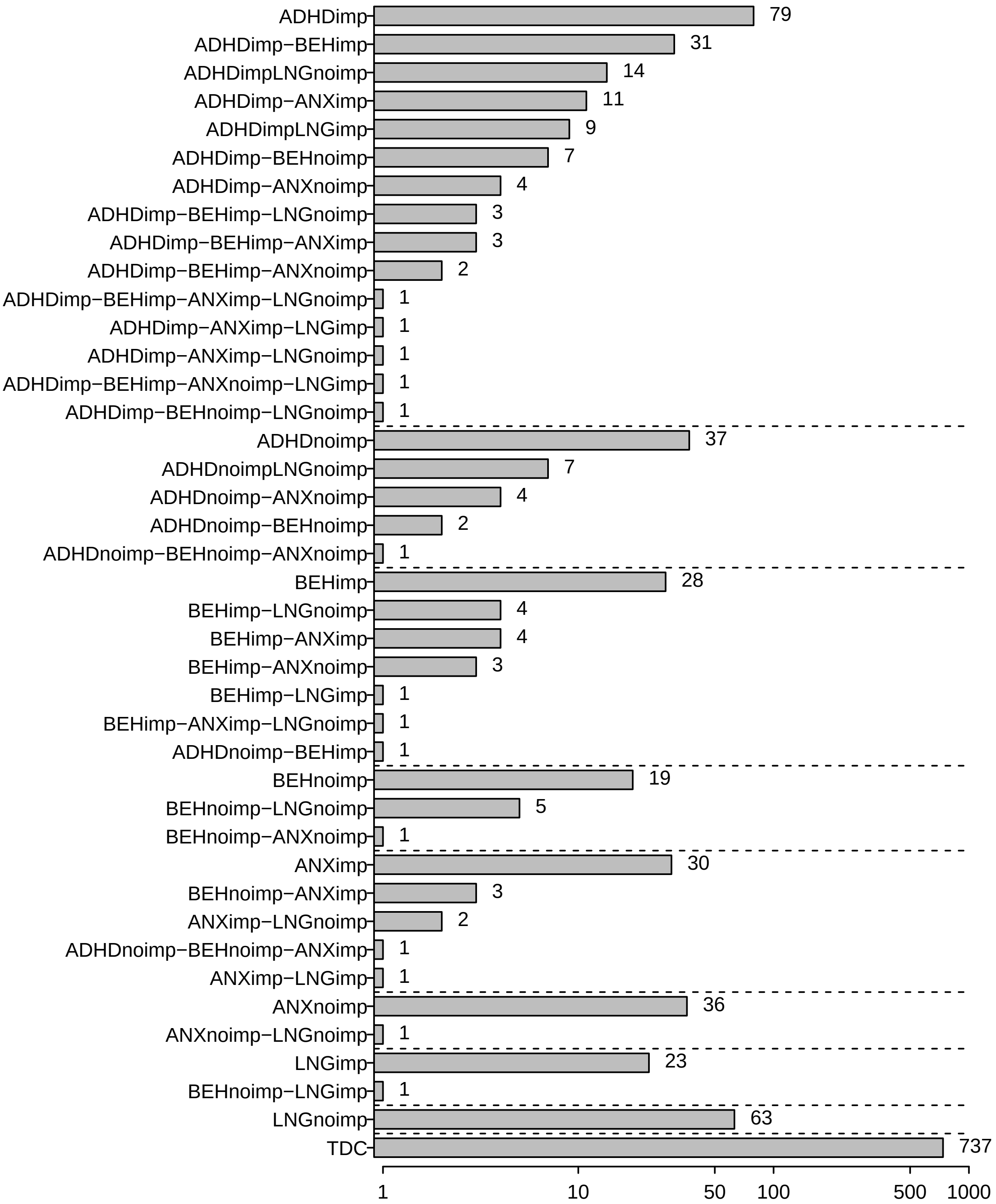


*Figure S3*. Combinations of mental health problems observed in the study. ADHD = Attention Deficit Hyperactivity Disorder, BEH = Behaviour problems, ANX = Anxiety problems, . . . imp = with impairment, . . . noimp = without impairment. Dotted horizontal lines delineate diagnostics groups used in the article.

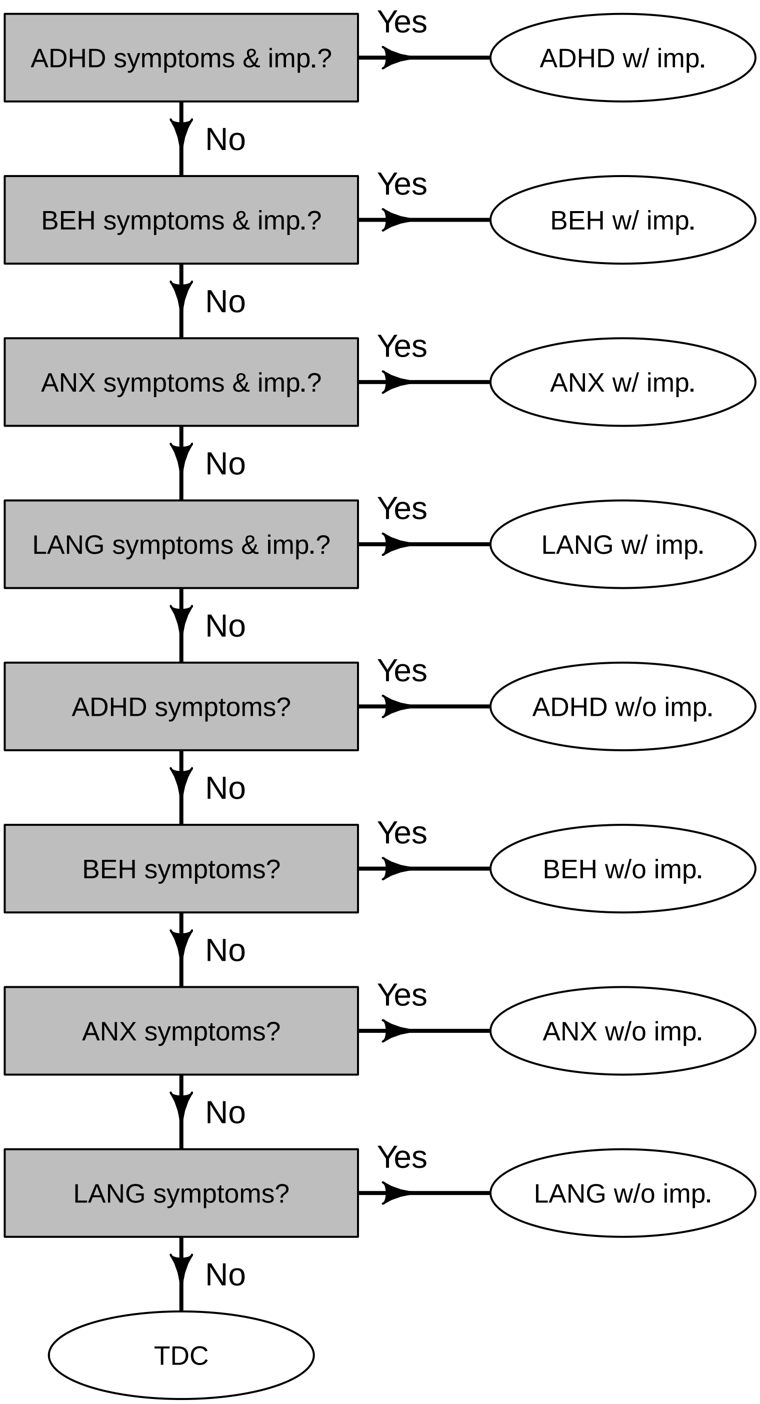


*Figure S4*. Decision tree for diagnostic classifications. For any mental health problems we first checked if children fulfilled all symptom and impairment criteria. Next we checked if children fulfilled symptom criteria. Children who did not fulfill any symptom criterion were classified as typically developing controls (TDC). ADHD = Attention Deficit Hyperactivity Disorder, BEH = Behaviour problems, ANX = Anxiety problems.

of an underlying normally distributed trait that determines item responses particularly lends itself to the estimation of approximately normally distributed scores for all scales. This in turn facilitates all further analyses. We assigned items to sub-scales as described in the respective manuals, and then estimated a Rasch model for each sub-scale (e.g. five Rasch models for the 5 BRIEF sub-scales).

**Testing the RDoC domain structure.** We used factor-analytic methods to identify a smaller number of functional dimensions from altogether 33 scales, comprised of sub-scale scores from questionnaires and tests. A first analysis used confirmatory factor analyses (CFA) to test if a CFA model with theoretically guided assignment of the scales to the RDoC dimensions captures the co-variation between test scores. We tested CFAs that used either

1. all available sub-scales

2. only sub-scales used for the final ESEM model (details below)

3. only sub-scales used for the final ESEM model which did not load on the introversion factor (see below)

4. as 3., and in addition sub-scales for the cognitive system were split into factor for questionnaire based and one for test based sub-scales.

The fit-indices Root Mean Square Error of Approximation (RMSEA) and Comparative Fit Index (CFI) shown in Table S1 indicate that these CFA models did not fit the data well. Hence we used exploratory structural equation modeling (ESEM, Asparouhov and Muthen, 2009), as implemented in the software Mplus, to identify functional domains in our data.

**Identification of functional domains through exploratory structural equation modeling (ESEM).** We used an ESEM analysis instead of a classical exploratory factor analysis (EFA) because it allows identification of correlated factors

Table S1

*Fit indices for RDoC CFAs. The third model builds on the second and the fourth on the third.*

| Model Description | CFI | TLI | RMSEA | ... 90%CI LB | ... 90%CI UB |
|---|---|---|---|---|---|
| all subscales | 0.454 | 0.408 | 0.081 | 0.079 | 0.084 |
| final ESEM sub-scales | 0.759 | 0.716 | 0.070 | 0.067 | 0.074 |
| ... + no introv. sub-scales | 0.537 | 0.468 | 0.096 | 0.093 | 0.100 |
| ... + 2 CS factors | 0.522 | 0.463 | 0.090 | 0.087 | 0.092 |

and extraction of factor scores in Mplus. Most sub-scales scores were treated as normally distributed, because we calculated standardized scores (e.g. for the Stanford Binet tests), or because Rasch models assume and typically produce (approximately) normally distributed person-scores. The few sub-scale score that were clearly not normally distributed (CDT, NEPSY visual attention), were categorized into 8 ordered scores and analyzed as ordered categorical data.

To deal with missing data, ESEM models were estimated with the Full Information Maximum Likelihood Method.

We used a 2-step approach to identify a set of functional domains. First, we fitted ESEM models with two to ten latent factors (dimensions/domains) to all sub-scales and chose the ESEM model with the smallest number of factors that had an RMSEA below 0.05 and CFI above 0.95 as the best model. Based on this model, we identified non-valid sub-scales as those that had a low $r^2$ value ($r^2 < 0.2$). The reasoning here is that that sub-scales with a very low $r^2$ value are either unreliable, or measure something that is unrelated to all other sub-scales. Using only the valid sub-scales we performed a second ESEM analysis. Based on the results of the second ESEM we identified ambiguous sub-scales as those with high cross-loadings

$\left(\frac{|highest\ loading|}{|2^{nd}\ highest\ loading|} < 1.25\right)$. Next we again fitted ESEM models with two to ten factors and identified the best model among these. Lastly, we investigated a possible simplification of the factor structure by testing CFA models in which low cross-loadings of scales were constrained to be zero.

**Comparison of functioning in preschoolers with and without ADHD.** We estimated a hierarchical regression model with the R package brms (Bürkner, 2017b). The basic regression model adjusts for maternal education and age, as well as for parity. The hierarchical parts of the model capture repeated measurement of individuals and model random effects for subsets of the data defined by by mental health problem, gender, and functional domain. The model also simultaneously imputed missing covariates. Specifically, the model was initially specified as follows:

effect_model = bf(value ~ 1 + mo(mEdu) + mo(parity) + Age + (1 | Domain:Gender:MHP) + (1|ID))

imputation_model = bf(mEdu ~ Age + poly(as.numeric(parity),2) + MHP, family = "cumulative")

joint_model = effect_model + imputation_model + set_rescor(FALSE)

fit = brm(joint_model, data = factor_scores_long, family = "gaussian", iter = 2000)

The model was fit with brms' default weakly informative priors. In particular, fixed effects and variance parameters have a student-t prior with mean zero, standard deviation 3, and 10 degrees of freedom. These priors lead to a, compared to uniform priors, weak shrinkage of parameter estimates towards zero. Due to the large sample size of the ADHD Study (~1200 participants), priors have a negligible effect on the results.

To verify successful convergence of the Bayesian estimation, we checked potential scale reduction parameters ($\hat{R}$) and divergent iterations. $\hat{R}$ for all random parameters were below 1.1 and the chains had no divergent iterations.

### Self selection into the Study

To assess self selection into the ADHD Study, we performed Bayesian multiple regression analyses to predict participation based on mothers' age and education (with the levels elementary school or less, high school, bachelor, or master degree), parity (0, 1, 2, 3 or more), and the child's sex and ADHD sum score in the MoBa Q6. For control participants we also used participation in the Q6 as a predictor. For this analysis, missing data were imputed during estimation of regression weights in a custom Stan (Carpenter *et al.*, 2017) program.

The results revealed some self selection in the ADHD Study. In the ADHD case group the chance to participate increased for each level of education, odds ratio (OR (95%) Highest Density Interval) 1.06 (1.04, 1.08), and increased substantially with each doubling of the ADHD sum score, OR 1.37 (1.23, 1.53). In the control group, the chance to participate was much lower for participants who had not returned the Q6, OR (95% Highest Density Interval, HDI) 0.68 (0.63, 0.72), ORs were 0.99 (0.96, 1.03) and 1.04 (0.99, 1.12) for education and ADHD scum score, respectively.

### Supplementary results: Factor loadings and correlations

Initial CFAs, for which sub-scales were on theoretical grounds assigned to RDoC domains, did not describe the data sufficiently well (RMSEA = 0.09 (0.087,0.092), CFI = 0.52, TLI = 0.46, see also Table S1 ). The first ESEM analysis including all 34 sub-scales showed that following scales either had an $r^2 < 0.2$ or high cross-loadings on a second factor and were thus removed from the analysis: BRIEF-Shift, CBQ-Discomfort, -Fear, and -Inhibitory control, NEPSY language test,

Stanford Binet's test for nonverbal IQ and verbal fluency. The final ESEM analysis with the remaining 26 sub-scales showed that a 7-factor model was the simplest model that could adequately describe the data (RMSEA = 0.03 (0.028, 0.037), CFI = 0.96). Follow up CFA analyses that constrained small cross loadings to 0 resulted in unsatisfactory RMSEA and CFI statistics. We hence retained the final ESEM model as the best model of functional domains for preschoolers in our sample.

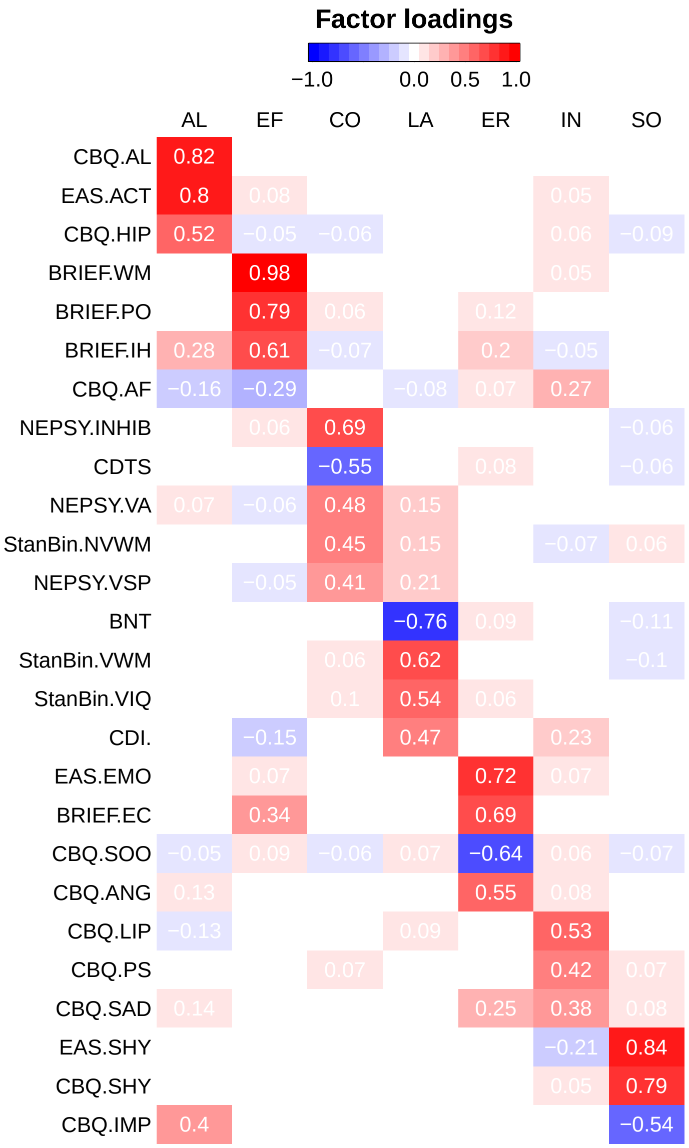


*Figure S5*. Factor loadings for the final ESEM model. AL = Activity level and regulation, EF = Executive functions, CO = cognition, LA = Language, ER = Emotion regulation, IN = Introversion, SO = Socialbility. See methods section for abbreviations of instruments and subscales.

Table S2

*x- and y- standardized factor loadings with standard errors*

| | AL | EF | CO | LA | ER | IN | SO |
|---|---|---|---|---|---|---|---|
| BNT | -0.04 (0.03) | -0.03 (0.03) | 0.03 (0.03) | -0.76 (0.04) | 0.09 (0.04) | 0.01 (0.03) | -0.11 (0.05) |
| BRIEF.EC | -0.04 (0.02) | 0.34 (0.04) | -0.00 (0.02) | 0.03 (0.02) | 0.69 (0.03) | -0.03 (0.01) | 0.01 (0.01) |
| BRIEF.IH | 0.28 (0.02) | 0.61 (0.02) | -0.07 (0.02) | 0.02 (0.02) | 0.20 (0.03) | -0.05 (0.02) | -0.01 (0.02) |
| BRIEF.PO | -0.00 (0.01) | 0.79 (0.02) | 0.06 (0.02) | -0.02 (0.02) | 0.12 (0.03) | -0.01 (0.02) | -0.02 (0.02) |
| BRIEF.WM | 0.01 (0.01) | 0.98 (0.01) | 0.01 (0.01) | -0.04 (0.02) | -0.03 (0.01) | 0.05 (0.02) | 0.03 (0.01) |
| CBQ.AF | -0.16 (0.04) | -0.29 (0.03) | 0.02 (0.03) | -0.08 (0.04) | 0.07 (0.03) | 0.27 (0.04) | -0.00 (0.03) |
| CBQ.AL | 0.82 (0.02) | 0.01 (0.01) | 0.03 (0.02) | -0.04 (0.02) | 0.02 (0.02) | -0.03 (0.02) | 0.01 (0.02) |
| CBQ.ANG | 0.13 (0.03) | 0.03 (0.02) | -0.04 (0.03) | -0.02 (0.03) | 0.55 (0.03) | 0.08 (0.03) | -0.03 (0.03) |
| CBQ.HIP | 0.52 (0.03) | -0.05 (0.03) | -0.06 (0.04) | 0.02 (0.03) | 0.02 (0.03) | 0.06 (0.03) | -0.09 (0.03) |
| CBQ.IMP | 0.40 (0.03) | -0.01 (0.02) | -0.02 (0.03) | 0.04 (0.03) | 0.01 (0.02) | -0.04 (0.02) | -0.54 (0.02) |
| CBQ.LIP | -0.13 (0.04) | -0.01 (0.02) | -0.04 (0.03) | 0.09 (0.04) | -0.03 (0.03) | 0.53 (0.04) | -0.03 (0.02) |
| CBQ.PS | 0.03 (0.02) | -0.03 (0.03) | 0.07 (0.05) | 0.03 (0.03) | -0.02 (0.03) | 0.42 (0.04) | 0.07 (0.04) |
| CBQ.SAD | 0.14 (0.04) | 0.00 (0.02) | 0.00 (0.03) | -0.04 (0.03) | 0.25 (0.04) | 0.38 (0.04) | 0.08 (0.03) |
| CBQ.SHY | 0.03 (0.02) | 0.03 (0.02) | -0.01 (0.02) | 0.03 (0.02) | 0.01 (0.02) | 0.05 (0.03) | 0.79 (0.02) |
| CBQ.SOO | -0.05 (0.02) | 0.09 (0.03) | -0.06 (0.04) | 0.07 (0.03) | -0.64 (0.03) | 0.06 (0.03) | -0.07 (0.03) |
| CDI. | 0.01 (0.02) | -0.15 (0.04) | 0.01 (0.03) | 0.47 (0.03) | 0.01 (0.02) | 0.23 (0.04) | -0.02 (0.02) |
| CDTS | 0.01 (0.03) | -0.03 (0.03) | -0.55 (0.05) | 0.03 (0.04) | 0.08 (0.04) | 0.02 (0.03) | -0.06 (0.04) |

Table S2 continued

| | AL | EF | CO | LA | ER | IN | SO |
|---|---|---|---|---|---|---|---|
| EAS.ACT | 0.80 (0.02) | 0.08 (0.02) | 0.01 (0.02) | -0.01 (0.02) | -0.04 (0.02) | 0.05 (0.02) | -0.02 (0.02) |
| EAS.EMO | -0.04 (0.02) | 0.07 (0.03) | -0.02 (0.03) | 0.03 (0.02) | 0.72 (0.03) | 0.07 (0.03) | -0.01 (0.02) |
| EAS.SHY | -0.04 (0.02) | -0.03 (0.02) | 0.00 (0.02) | 0.00 (0.01) | 0.03 (0.02) | -0.21 (0.04) | 0.84 (0.03) |
| NEPSY.INHIB | -0.02 (0.03) | 0.06 (0.03) | 0.69 (0.04) | -0.02 (0.03) | 0.00 (0.02) | 0.04 (0.03) | -0.06 (0.03) |
| NEPSY.VA | 0.07 (0.04) | -0.07 (0.04) | 0.48 (0.05) | 0.15 (0.05) | 0.04 (0.04) | 0.00 (0.04) | 0.01 (0.03) |
| NEPSY.VSP | -0.03 (0.03) | -0.05 (0.03) | 0.41 (0.04) | 0.21 (0.05) | 0.04 (0.03) | 0.03 (0.04) | -0.03 (0.03) |
| StanBin.NVWM | 0.00 (0.03) | -0.02 (0.03) | 0.45 (0.05) | 0.15 (0.05) | -0.04 (0.03) | -0.07 (0.04) | 0.06 (0.03) |
| StanBin.VIQ | -0.04 (0.03) | -0.01 (0.03) | 0.10 (0.05) | 0.55 (0.04) | 0.06 (0.03) | 0.01 (0.03) | -0.01 (0.02) |
| StanBin.VWM | -0.03 (0.03) | 0.01 (0.03) | 0.06 (0.04) | 0.62 (0.04) | 0.01 (0.02) | -0.02 (0.03) | -0.10 (0.04) |

*Note.* Numbers are mean and confidence intervals. AL = Activity level and regulation, EF = Executive functions, CO = cognition, LA = Language, ER = Emotion regulation, IN = Introversion, SO = Socialbility. See methods section for abbreviations of instruments and subscales.

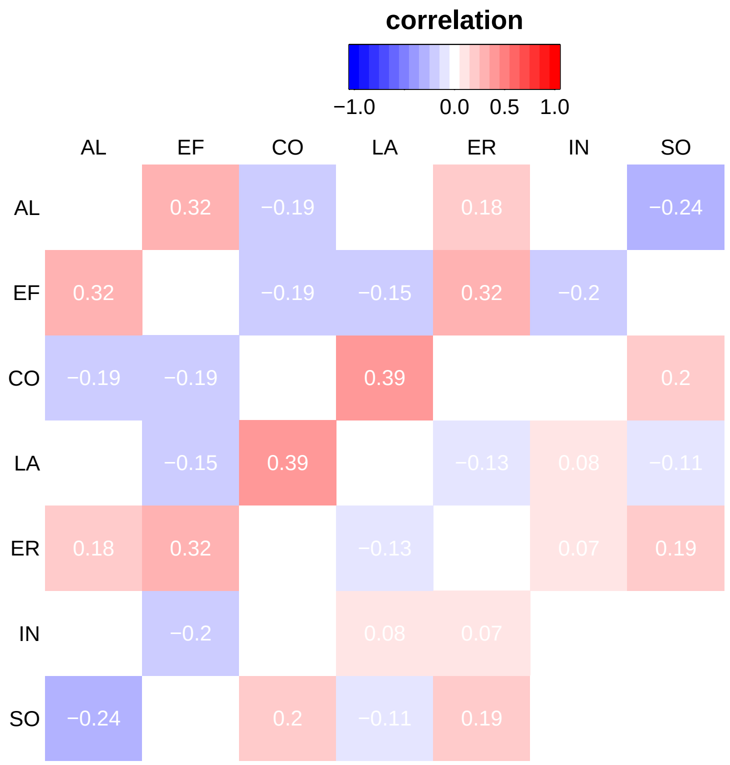


*Figure S6*. Factor correlations for the final ESEM model. AL = Activity level and regulation, EF = Executive functions, CO = cognition, LA = Language, ER = Emotion regulation, IN = Introversion, SO = Socialbility.

Table S3

*Factor correlations with confidence intervals*

| | EF | CO | LA | ER | IN | SO |
|---|---|---|---|---|---|---|
| AL | 0.33 (-0.31, 0.96) | 0.32 (-0.31, 0.96) | -0.15 ( 0.14, -0.45) | 0.03 (-0.03, 0.08) | -0.19 ( 0.19, -0.57) | -0.20 ( 0.19, -0.59) |
| EF | | 0.18 (-0.17, 0.53) | -0.13 ( 0.12, -0.37) | 0.19 (-0.18, 0.56) | -0.01 ( 0.01, -0.03) | 0.07 (-0.07, 0.20) |
| CO | | | -0.04 ( 0.04, -0.11) | -0.24 ( 0.23, -0.72) | -0.19 ( 0.18, -0.56) | -0.02 ( 0.02, -0.06) |
| LA | | | | -0.11 ( 0.10, -0.31) | 0.39 (-0.37, 1.15) | 0.08 (-0.08, 0.24) |
| ER | | | | | 0.20 (-0.20, 0.60) | 0.04 (-0.04, 0.13) |
| IN | | | | | | 0.03 (-0.03, 0.09) |

*Note.* Numbers are means and confidence intervals. AL = Activity level and regulation, EF = Executive functions, CO = cognition, LA = Language, ER = Emotion regulation, IN = Introversion, SO = Socialbility.

Table S4

*Average differences in functioning across domains between preschoolers with mental health problems and typically developing preschoolers*

| | with impairment | without impairment |
|---|---|---|
| ADHD | -0.55 (-0.62, -0.48; 0.91) | -0.28 (-0.40, -0.17; 0.00) |
| BEH | -0.33 (-0.45, -0.21; 0.01) | -0.29 (-0.44, -0.15; 0.00) |
| ANX | -0.20 (-0.33, -0.08; 0.00) | -0.17 (-0.29, -0.04; 0.00) |
| LNG | -0.52 (-0.66, -0.36; 0.60) | -0.30 (-0.41, -0.20; 0.00) |

*Note.* Standardized mean deviations (SMD). Values are mean (lower 90% HDI, upper 90% HDI; P(SMD<-0.5)). ADHD = preschoolers with AHDH problems, BEH = behavior problems, ANX = anxiety, LNG = language.

**Supplementary results: Group comparisons**

Table S5

*Differences in functioning between preschoolers with ADHD and typically developing preschoolers, stratified by gender or severity of mental health problem.*

| | V1 | with impairment | without impairment | boy | girl |
|---|---|---|---|---|---|
| AL | -0.92 (-1.07, -0.78; 1.00) | -0.97 (-1.14, -0.82; 1.00) | -0.74 (-0.99, -0.47; 0.97) | -0.80 (-1.01, -0.57; 1.00) | -0.92 (-1.14, -0.70; 1.00) |
| EF | -0.91 (-1.04, -0.77; 1.00) | -1.02 (-1.17, -0.87; 1.00) | -0.58 (-0.82, -0.33; 0.74) | -0.73 (-0.93, -0.51; 0.98) | -0.86 (-1.07, -0.65; 1.00) |
| CO | -0.51 (-0.65, -0.37; 0.58) | -0.55 (-0.70, -0.40; 0.73) | -0.40 (-0.66, -0.17; 0.22) | -0.46 (-0.67, -0.25; 0.37) | -0.49 (-0.70, -0.26; 0.45) |
| LA | -0.43 (-0.55, -0.28; 0.16) | -0.53 (-0.68, -0.38; 0.67) | -0.09 (-0.34, 0.15; 0.00) | -0.28 (-0.49, -0.07; 0.02) | -0.34 (-0.55, -0.12; 0.08) |
| ER | -0.62 (-0.75, -0.47; 0.95) | -0.71 (-0.86, -0.55; 0.99) | -0.31 (-0.56, -0.06; 0.07) | -0.44 (-0.65, -0.23; 0.30) | -0.58 (-0.80, -0.36; 0.75) |
| IN | -0.11 (-0.24, 0.04; 0.00) | -0.18 (-0.32, -0.01; 0.00) | 0.12 (-0.13, 0.36; 0.00) | -0.12 (-0.32, 0.11; 0.00) | 0.06 (-0.17, 0.27; 0.00) |
| SO | 0.09 (-0.07, 0.22; 0.00) | 0.11 (-0.05, 0.27; 0.00) | 0.02 (-0.21, 0.26; 0.00) | -0.14 (-0.35, 0.08; 0.00) | 0.26 ( 0.05, 0.49; 0.00) |

*Note.* AL = Activity level and regulation, EF = Executive functions, CO = cognition, LA = Language, ER = Emotion regulation, IN = Introversion, SO = Socialbility. Values are standardized mean deviations (lower and upper 90% credible interval; P(SMD<-0.5))

### Comparison of ADHD and ABC-Study samples

The sample of the ADHD study does not include children with ADS, because these children were recruited into ABC Study, a sister study about ASD. The following plots show data for tests and questionnaires that were available for both study (for the ABC Study, only around 50 of the full samples has data from the CBQ and EAS).

Children in the ABC study were classified into groups following the same algorithm described above. For the following analysis, each group with a mental health problems includes children with clinical and sub-clinical[2] classification.

The following figures show largely consistent results for the comparison of children with an without ADHD. One exception are children with language problems: Children with language problems in the ABC Study have greater language related problems than children with language problems in the ADHD study. This is likely explained by the fact that the ABC's studies attempt to recruit children with ASD also resulted in recruitment of children with sever language problems.

[2] i.e. they fullfull symptom-criteria but have only limited impairments, or they have clear impairments despite not fulfilling symptom criteria

Table S6

*Differences in functioning between preschoolers with ADHD and other mental health problems.*

| Comparison Group | Domain | ADHD w/ impairm. | ADHD w/o impairm. |
|---|---|---|---|
| ANX w/ impairm. | AL | -1.29 (-1.57, -0.98; 1.00) | -1.06 (-1.43, -0.71; 1.00) |
| | EF | -0.84 (-1.13, -0.52; 0.98) | -0.39 (-0.75, -0.04; 0.28) |
| | CO | -0.72 (-1.02, -0.42; 0.93) | -0.58 (-0.92, -0.20; 0.67) |
| | LA | -0.30 (-0.61, 0.01; 0.11) | 0.14 (-0.20, 0.51; 0.00) |
| | ER | -0.17 (-0.46, 0.13; 0.01) | 0.23 (-0.13, 0.57; 0.00) |
| | IN | -0.24 (-0.57, 0.06; 0.05) | 0.05 (-0.32, 0.39; 0.00) |
| | SO | 1.12 ( 0.82, 1.42; 0.00) | 1.03 ( 0.66, 1.36; 0.00) |
| BEH w/ impairm. | AL | -0.73 (-1.02, -0.44; 0.94) | -0.50 (-0.85, -0.15; 0.49) |
| | EF | -0.56 (-0.86, -0.27; 0.65) | -0.12 (-0.45, 0.22; 0.01) |
| | CO | -0.44 (-0.72, -0.15; 0.34) | -0.29 (-0.65, 0.04; 0.12) |
| | LA | -0.15 (-0.44, 0.14; 0.01) | 0.30 (-0.04, 0.65; 0.00) |
| | ER | 0.18 (-0.11, 0.47; 0.00) | 0.58 ( 0.23, 0.90; 0.00) |
| | IN | -0.08 (-0.36, 0.21; 0.00) | 0.21 (-0.15, 0.55; 0.00) |
| | SO | 0.26 (-0.03, 0.55; 0.00) | 0.17 (-0.16, 0.52; 0.00) |
| LNG w/ impairm. | AL | -1.11 (-1.49, -0.76; 1.00) | -0.88 (-1.28, -0.48; 0.97) |
| | EF | -0.42 (-0.82, -0.08; 0.34) | 0.02 (-0.42, 0.41; 0.01) |
| | CO | 0.21 (-0.17, 0.56; 0.00) | 0.36 (-0.04, 0.77; 0.00) |
| | LA | 1.20 ( 0.83, 1.59; 0.00) | 1.65 ( 1.25, 2.11; 0.00) |
| | ER | -0.52 (-0.87, -0.14; 0.54) | -0.12 (-0.52, 0.28; 0.03) |
| | IN | 0.32 (-0.05, 0.66; 0.00) | 0.61 ( 0.21, 1.02; 0.00) |
| | SO | 0.10 (-0.24, 0.45; 0.00) | 0.01 (-0.38, 0.41; 0.01) |

*Note.* AL = Activity level and regulation, EF = Executive functions, CO = cognition, LA = Language, ER = Emotion regulation, IN = Introversion, SO = Socialbility. Values are standardized mean deviations (lower and upper 90% credible interval; P(SMD<-0.5))

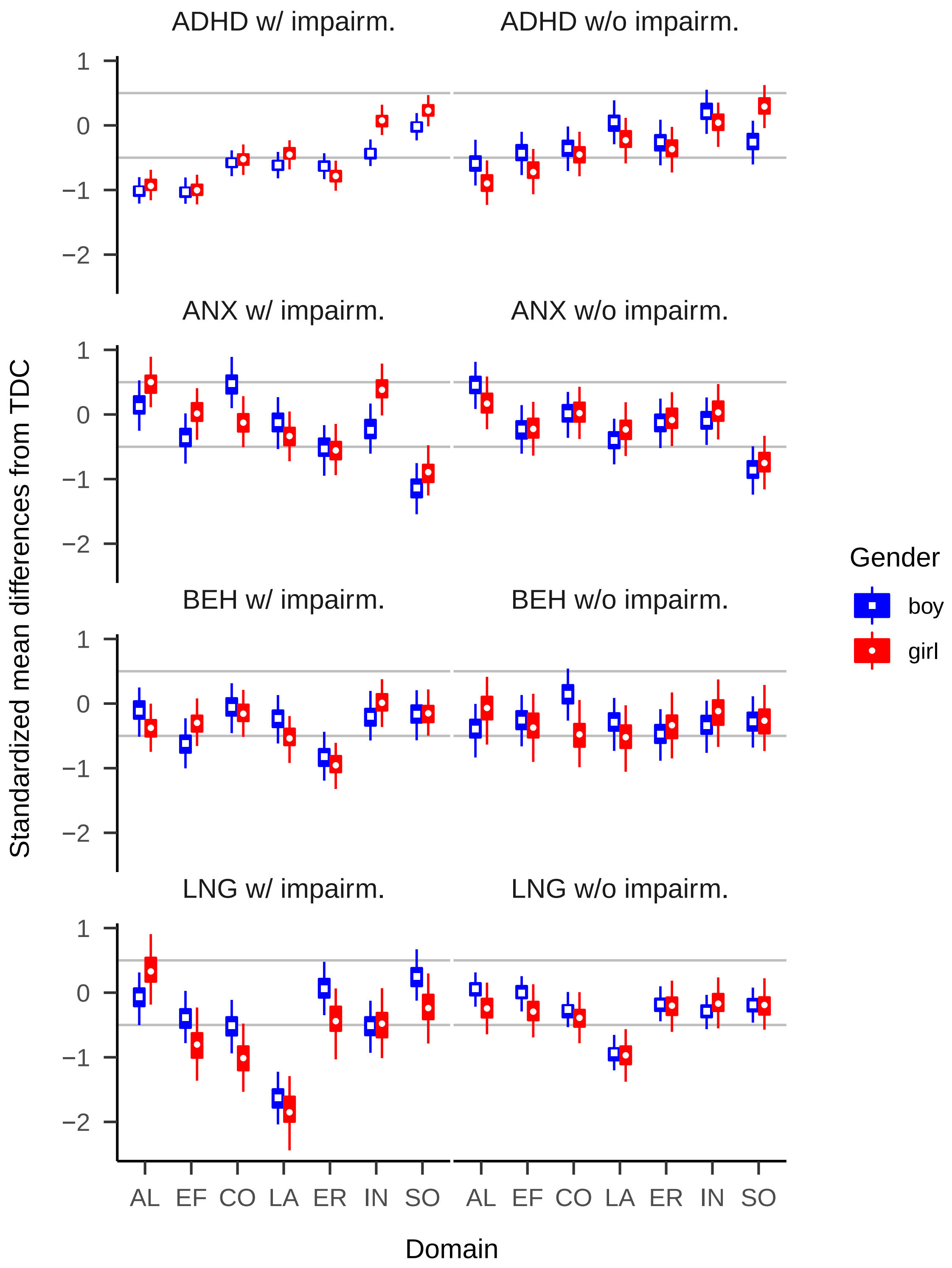


*Figure S7.* Comparison of children with mental health problems against typically developing controls, stratified by mental health problem type and gender. AL = Activity level and regulation, EF = Executive functions, CO = cognition, LA = Language, ER = Emotion regulation, IN = Introversion, SO = Socialbility. ADHD = preschoolers with AHDH problems, BEH = behavior problems, ANX = anxiety, LNG = language.

Table S7

*Differences in functioning between preschoolers with ADHD and other mental health problems by domain and severity.*

| Comparison Group | Domain | w/ impairm. | w/o impairm. |
|---|---|---|---|
| ALL | AL | -1.04 (-1.47, -0.57; 0.98) | -0.74 (-1.25, -0.29; 0.78) |
| | EF | -0.61 (-1, -0.22; 0.66) | -0.35 (-0.66, -0.01; 0.22) |
| | CO | -0.32 (-0.89, 0.4; 0.42) | -0.24 (-0.63, 0.13; 0.14) |
| | LA | 0.25 (-0.48, 1.41; 0.04) | 0.47 (-0.01, 1.04; 0) |
| | ER | -0.17 (-0.71, 0.34; 0.18) | -0.08 (-0.43, 0.31; 0.02) |
| | IN | 0 (-0.41, 0.51; 0.02) | 0.28 (-0.07, 0.62; 0) |
| | SO | 0.49 (-0.09, 1.28; 0) | 0.44 (-0.02, 1.01; 0) |
| BEH | AL | -0.73 (-0.97, -0.48; 0.94) | -0.51 (-0.84, -0.19; 0.52) |
| | EF | -0.56 (-0.8, -0.31; 0.65) | -0.26 (-0.61, 0.08; 0.13) |
| | CO | -0.44 (-0.68, -0.19; 0.34) | -0.24 (-0.57, 0.11; 0.09) |
| | LA | -0.15 (-0.39, 0.09; 0.01) | 0.32 (-0.02, 0.66; 0) |
| | ER | 0.18 (-0.06, 0.43; 0) | 0.09 (-0.24, 0.43; 0) |
| | IN | -0.08 (-0.33, 0.16; 0) | 0.34 (0.01, 0.68; 0) |
| | SO | 0.26 (0.02, 0.51; 0) | 0.29 (-0.05, 0.62; 0) |
| ANX | AL | -1.29 (-1.54, -1.03; 1) | -1.05 (-1.35, -0.76; 1) |
| | EF | -0.84 (-1.09, -0.58; 0.98) | -0.35 (-0.65, -0.05; 0.21) |
| | CO | -0.72 (-0.97, -0.47; 0.93) | -0.42 (-0.72, -0.12; 0.33) |
| | LA | -0.3 (-0.57, -0.04; 0.11) | 0.23 (-0.06, 0.52; 0) |
| | ER | -0.17 (-0.42, 0.08; 0.01) | -0.2 (-0.52, 0.1; 0.06) |
| | IN | -0.24 (-0.51, 0.02; 0.05) | 0.15 (-0.15, 0.46; 0) |
| | SO | 1.12 (0.87, 1.38; 0) | 0.82 (0.52, 1.13; 0) |
| LNG | AL | -1.11 (-1.42, -0.81; 1) | -0.65 (-0.94, -0.36; 0.81) |
| | EF | -0.42 (-0.73, -0.12; 0.34) | -0.43 (-0.7, -0.15; 0.34) |
| | CO | 0.21 (-0.08, 0.52; 0) | -0.08 (-0.36, 0.21; 0.01) |
| | LA | 1.2 (0.89, 1.53; 0) | 0.86 (0.58, 1.14; 0) |
| | ER | -0.52 (-0.82, -0.21; 0.54) | -0.12 (-0.4, 0.16; 0.01) |
| | IN | 0.32 (0.01, 0.62; 0) | 0.35 (0.07, 0.62; 0) |
| | SO | 0.1 (-0.19, 0.39; 0) | 0.21 (-0.06, 0.47; 0) |

*Note.* Values are standardized mean deviations (lower and upper 90% credible interval; P(SMD<-0.5)). ADHD = preschoolers with AHDH problems, BEH = behavior problems, ANX = anxiety, LNG = language. AL = Activity level and regulation, EF = Executive functions, CO = cognition, LA = Language, ER = Emotion regulation, IN = Introversion, SO = Socialbility.

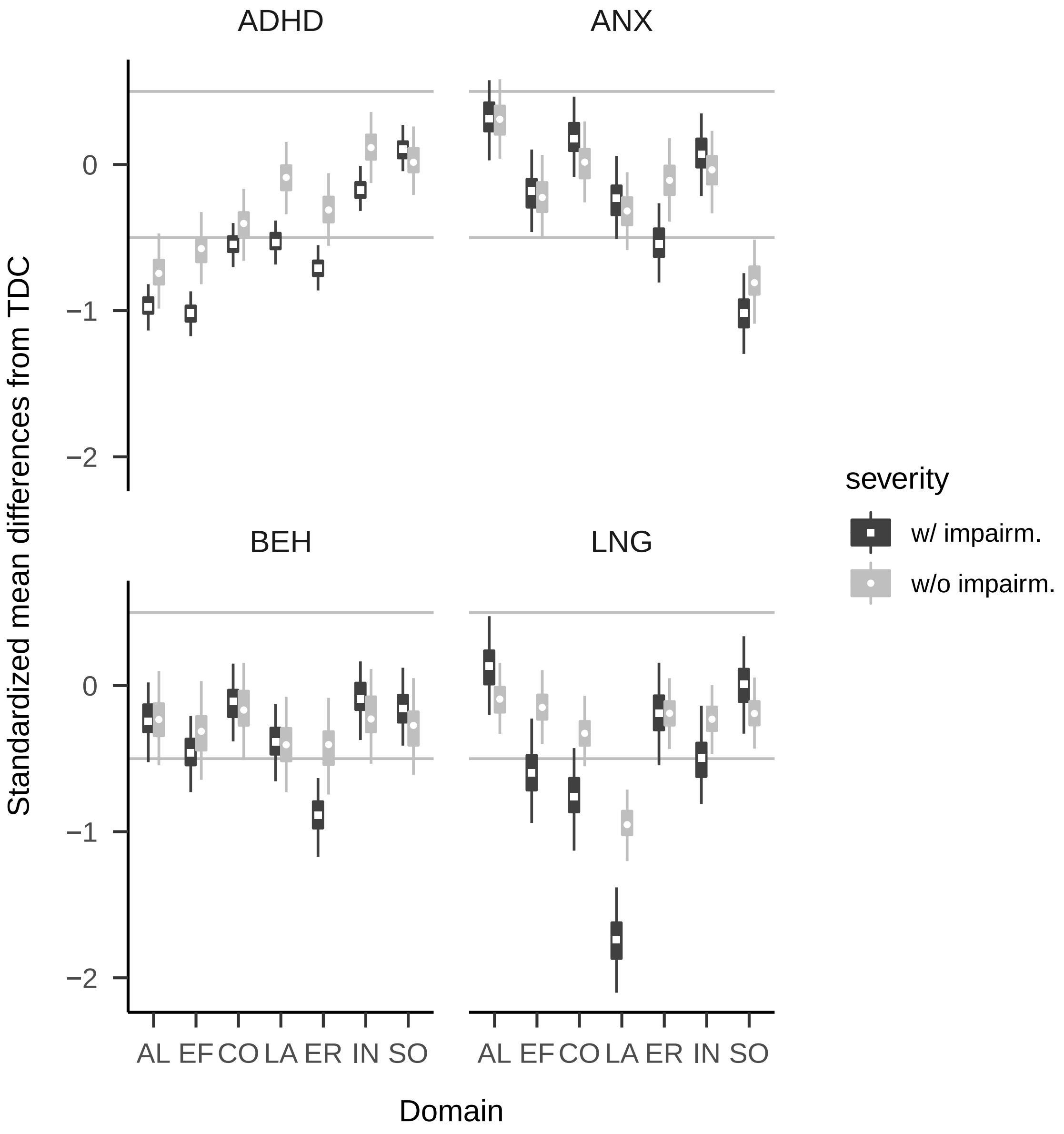


*Figure S8*. Comparison of children with mental health problems against typically developing controls, stratified by mental health problem type and problem severity (clinical and sub-clinical). AL = Activity level and regulation, EF = Executive functions, CO = cognition, LA = Language, ER = Emotion regulation, IN = Introversion, SO = Socialbility. ADHD = preschoolers with AHDH problems, BEH = behavior problems, ANX = anxiety, LNG = language.

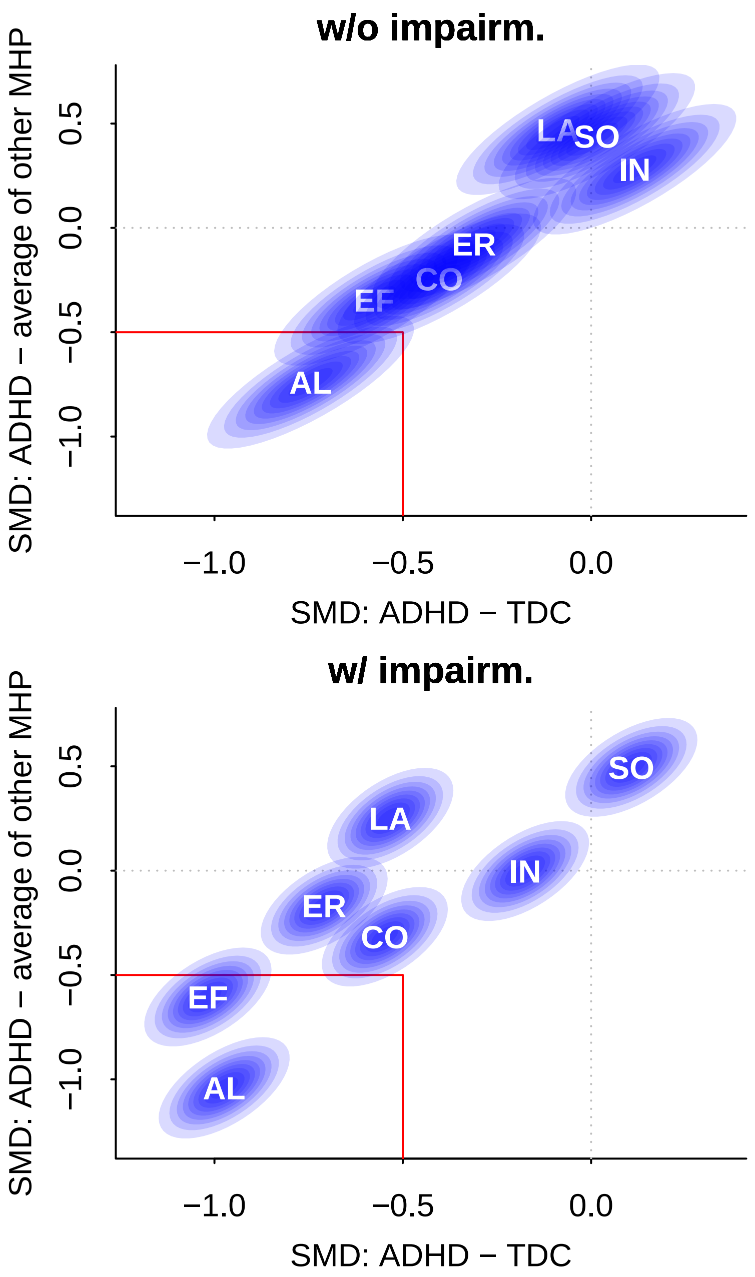


*Figure S9*. Comparison of preschoolers with ADHD with typically developing preschoolers and preschoolers with other *clinical* mental health problems, stratified by severity of impairment. Values below 0 indicate that the comparison group has fewer problems than preschoolers with ADHD. Ellipses cover 90% HDIs. Labels in ellipses indicate functional domains. Domains below the diagonal line are those where the sum of the SMD differences to TDCs and other mental health problems is 1. Domains in the lower left rectangle are those where the SMD differences to both TDCs and other mental health problems is at least 0.5.

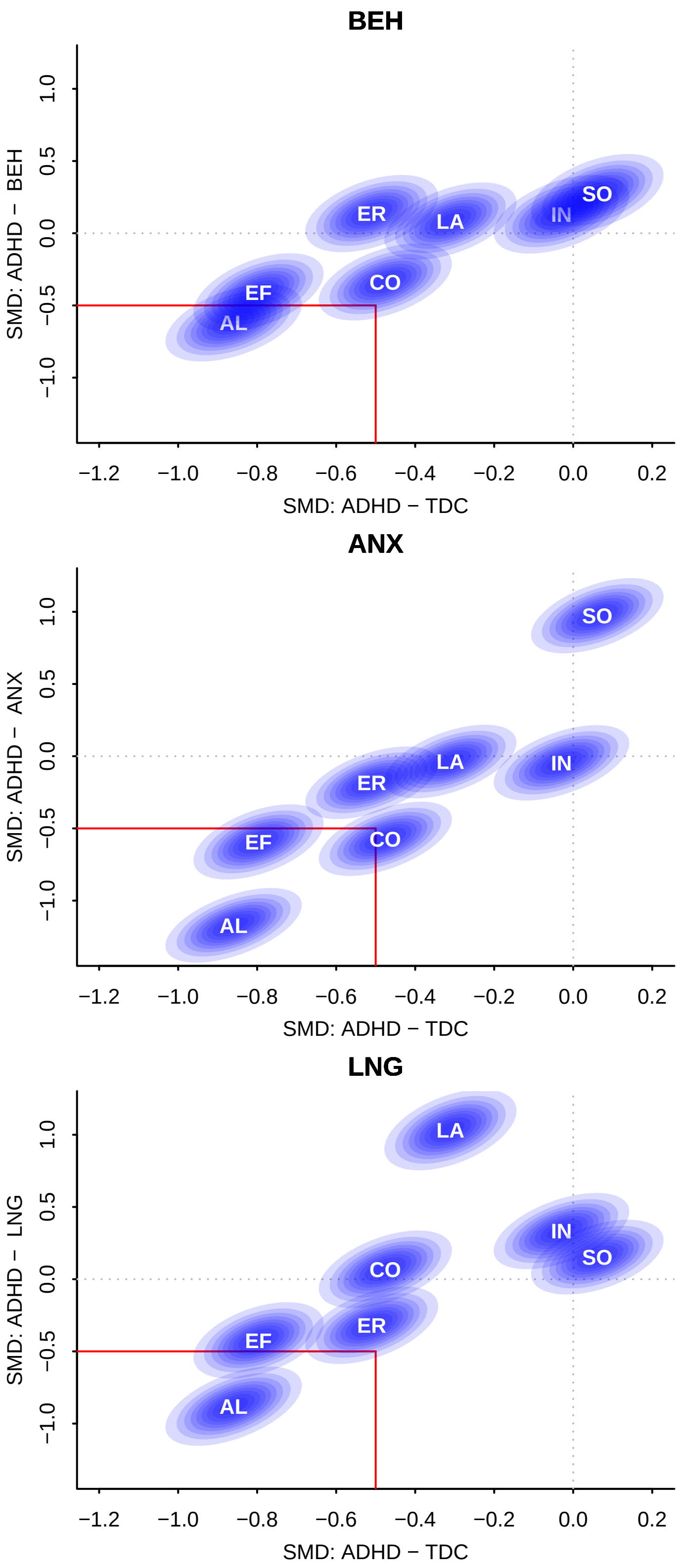


*Figure S10*. Comparison of preschoolers with ADHD with typically developing preschoolers and preschoolers with other *clinical* mental health problems.

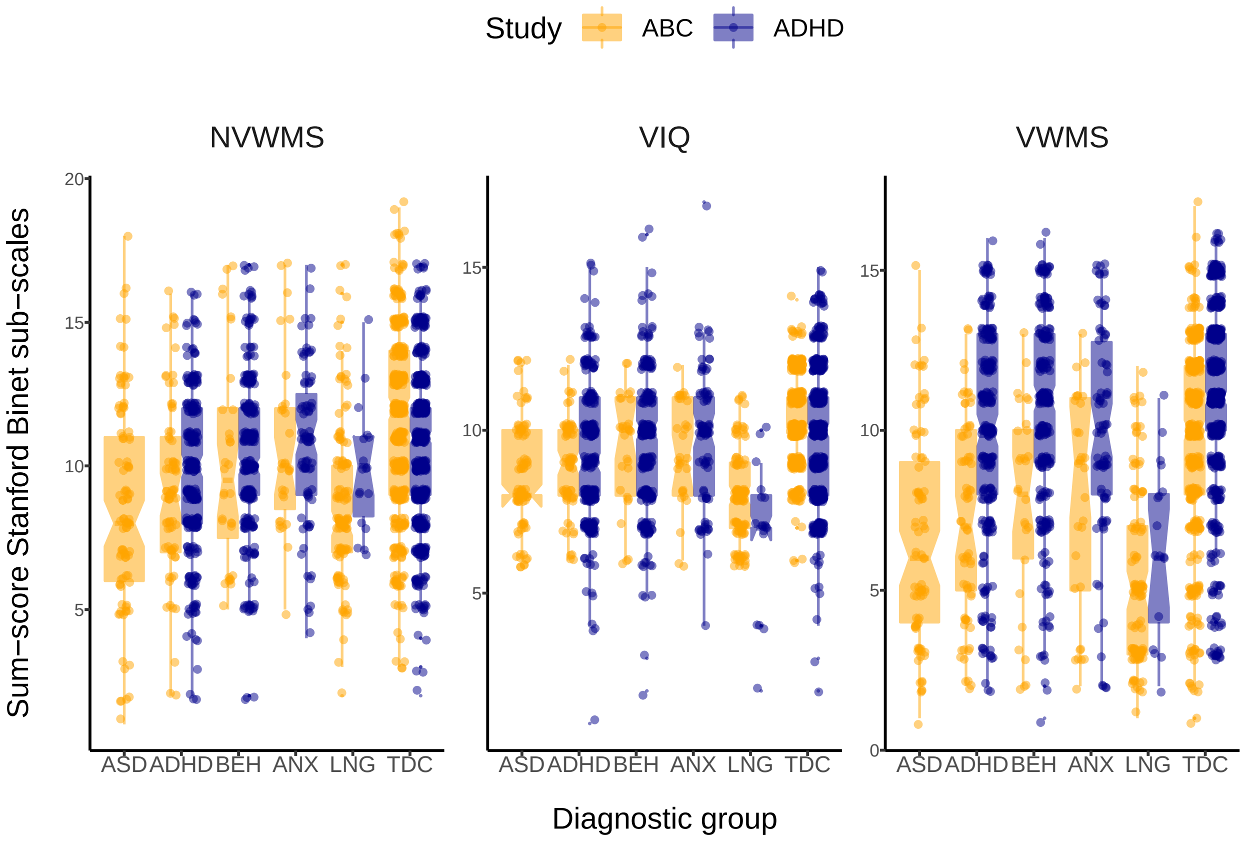


*Figure S11.* Stanford Binet Test results in the ADHD and ABC studies. Individual data points (dots) and boxplots with median, 25% and 75% quatiles. Notches, which are calculated as 1.58 * interquartile range divided by the squareroot of n, approximate 95% confidence intervals.

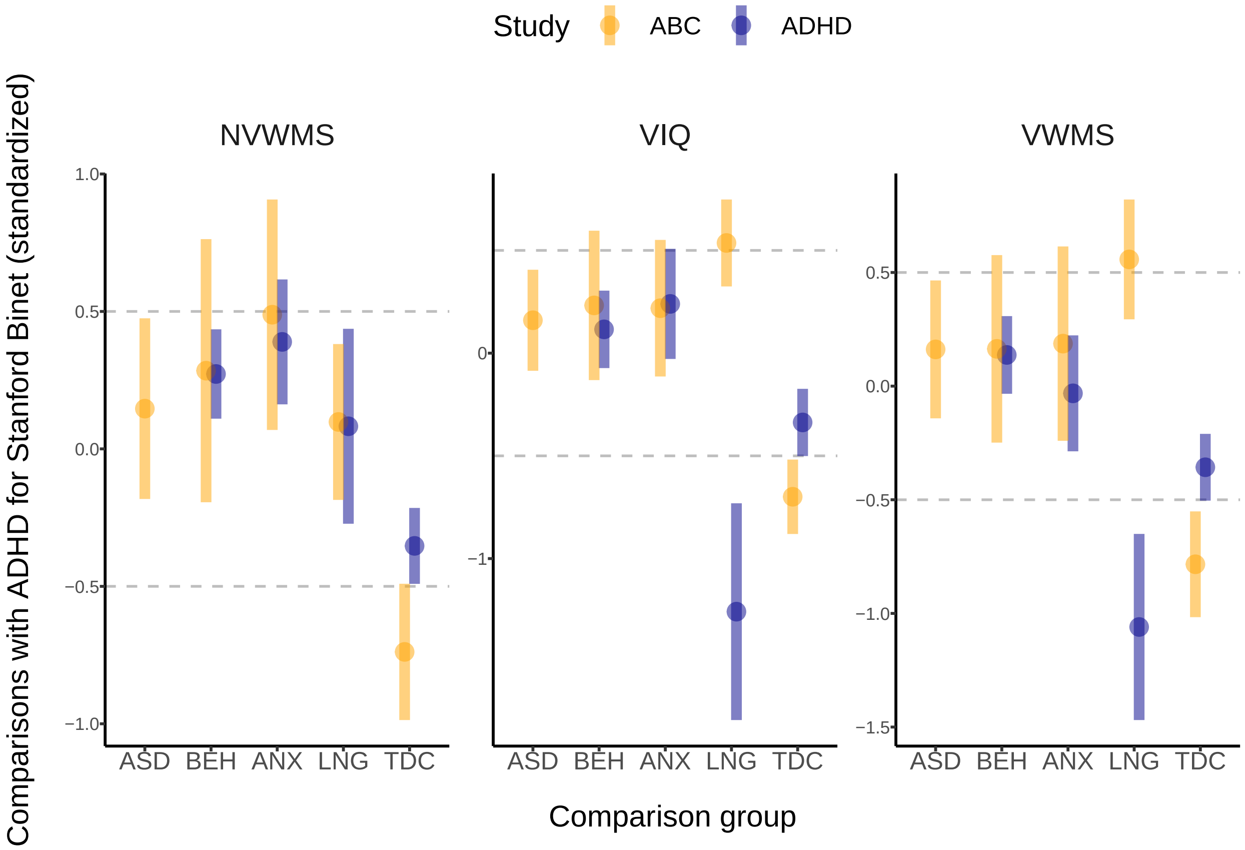


*Figure S12.* Comparison of children from ADHD and ABC Studies across Stanford Binet Test results. All differences are based on scaled scores, whereby scaling was implemented by dividing the raw scores by the standardeviation of all scores (across sub-groups and studies).

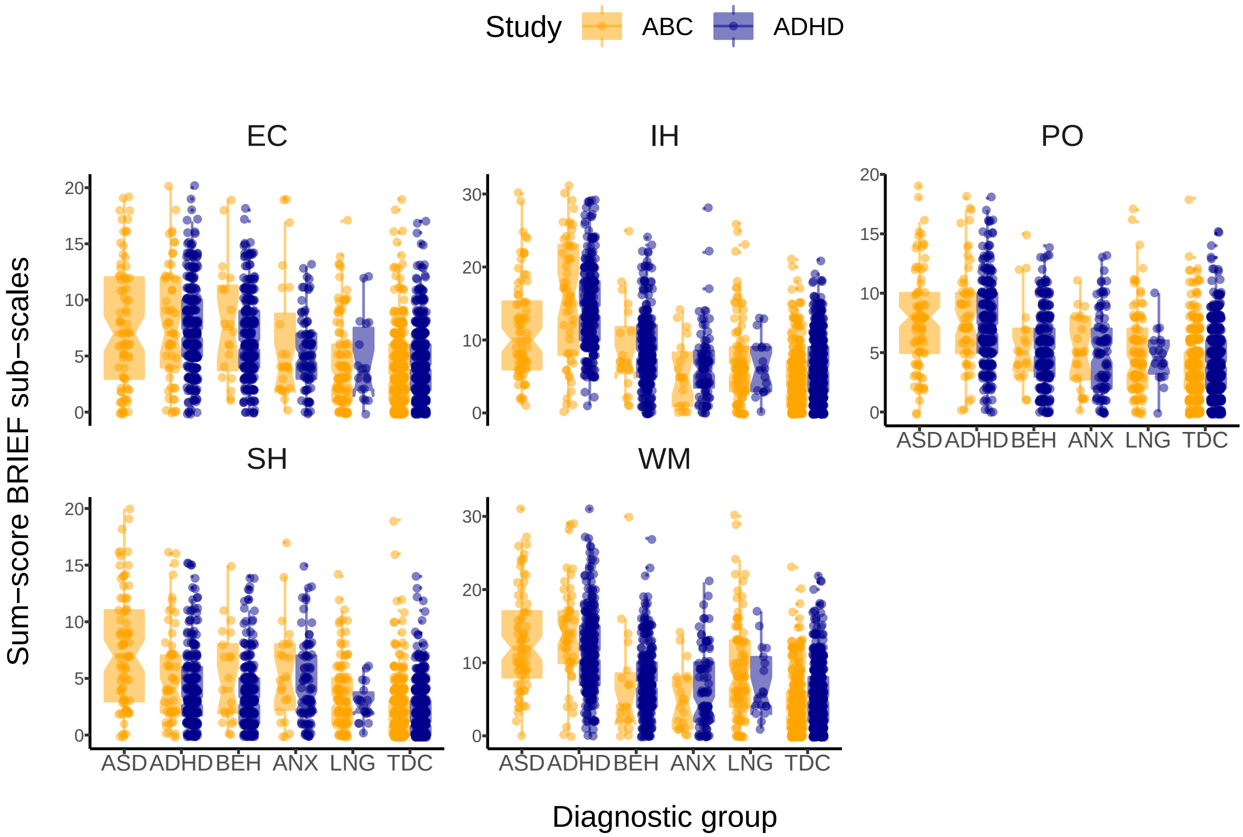


*Figure S13*. BRIEF results in the ADHD and ABC studies, stratified by group.

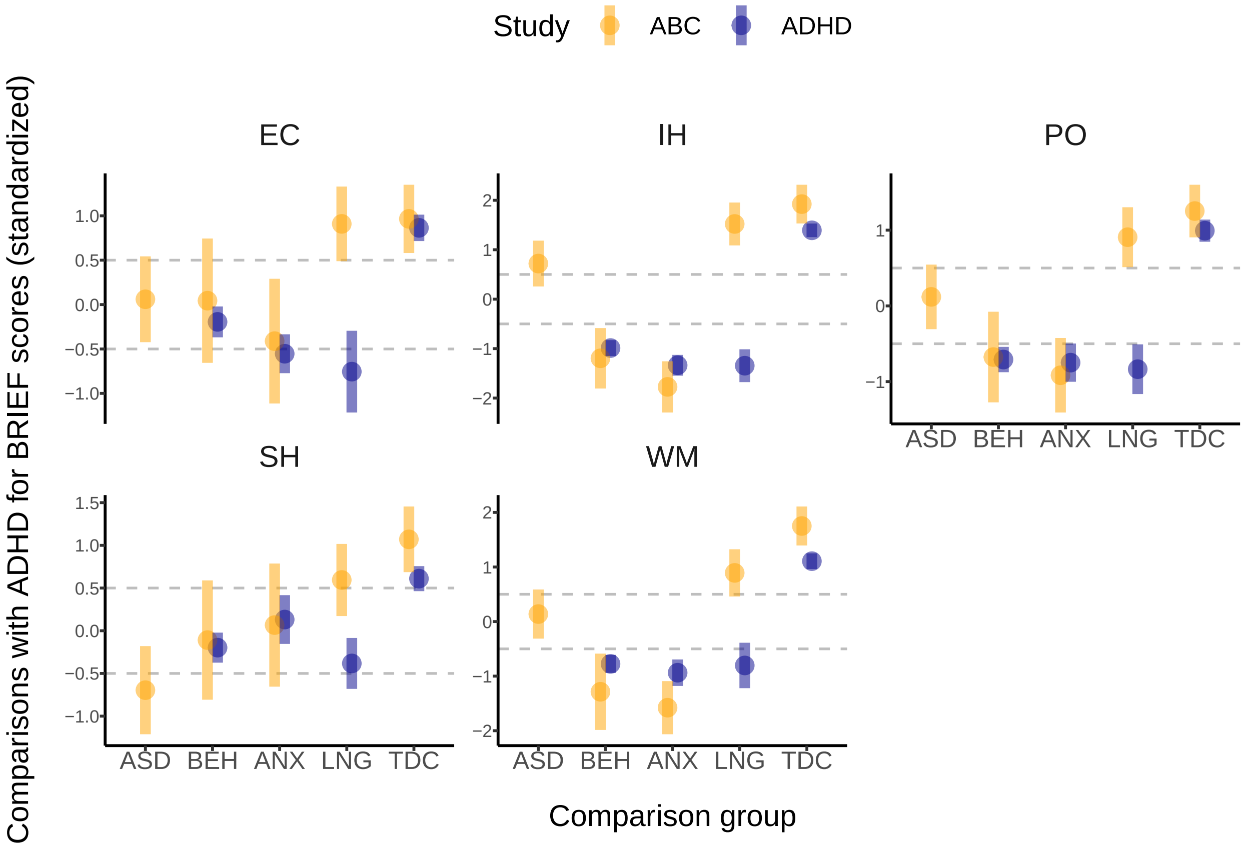


*Figure S14.* Comparison of children from ADHD and ABC Studies across BRIEF results. All differences are based on scaled scores, whereby scaling was implemented by deviding the raw scores by the standardeviation of all scores (across sub-groups and studies).

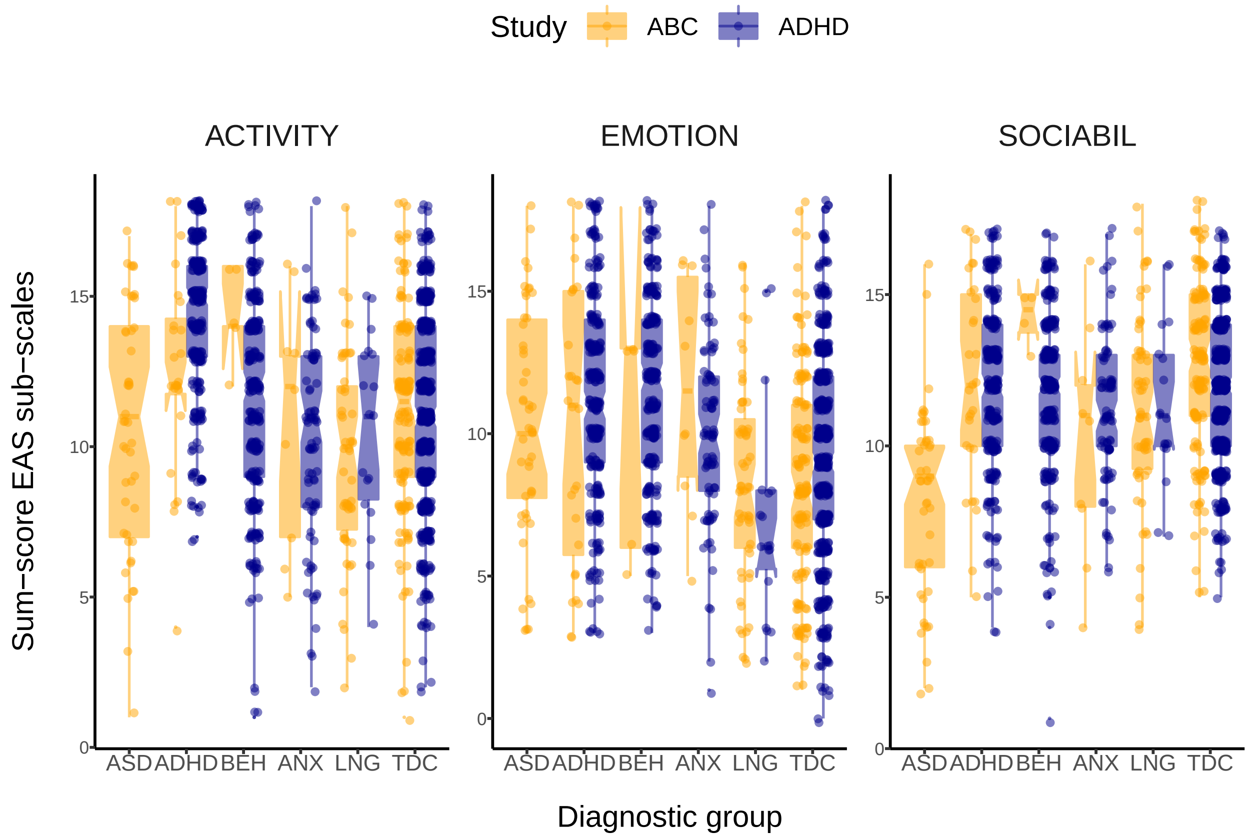


*Figure S15*. EAS results in the ADHD and ABC studies, stratified by group.

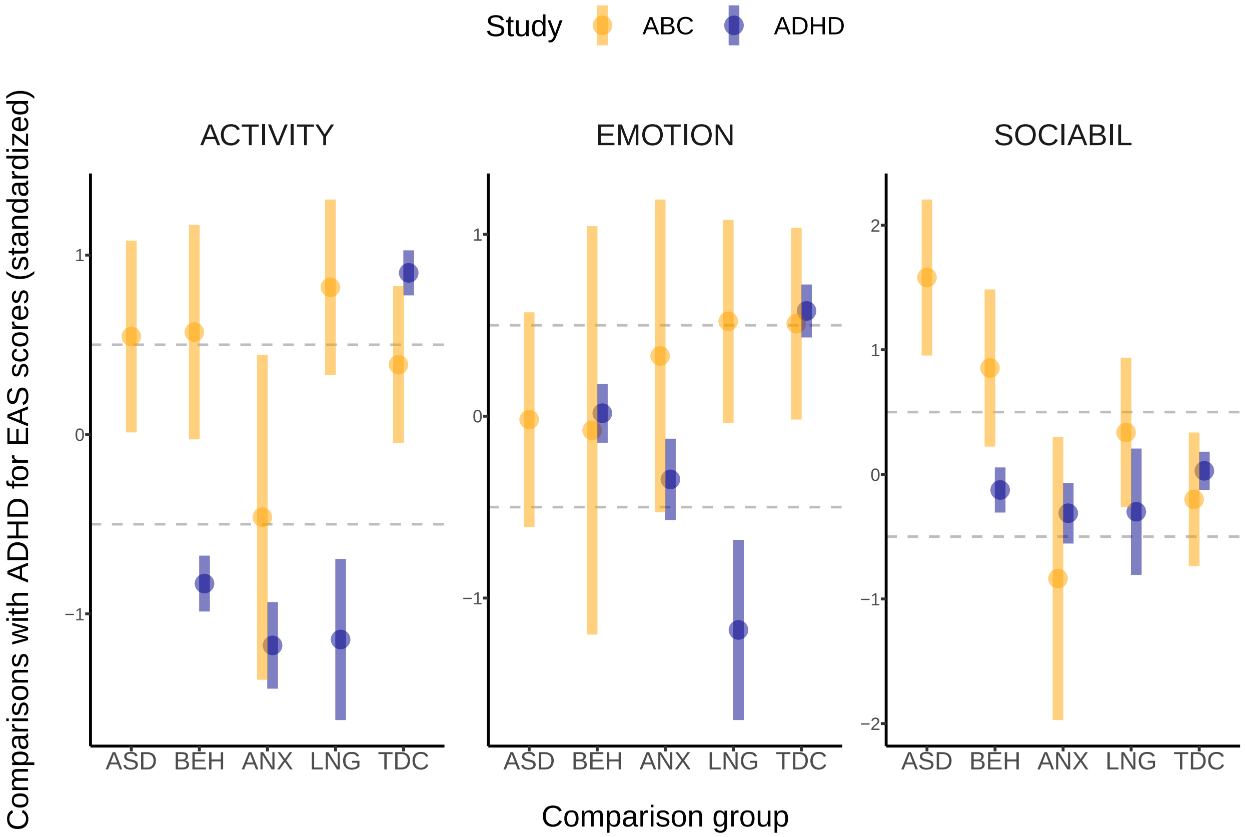


*Figure S16*. Comparison of children from ADHD and ABC Studies across EAS results. All differences are based on scaled scores, whereby scaling was implemented by deviding the raw scores by the standardeviation of all scores (across sub-groups and studies).

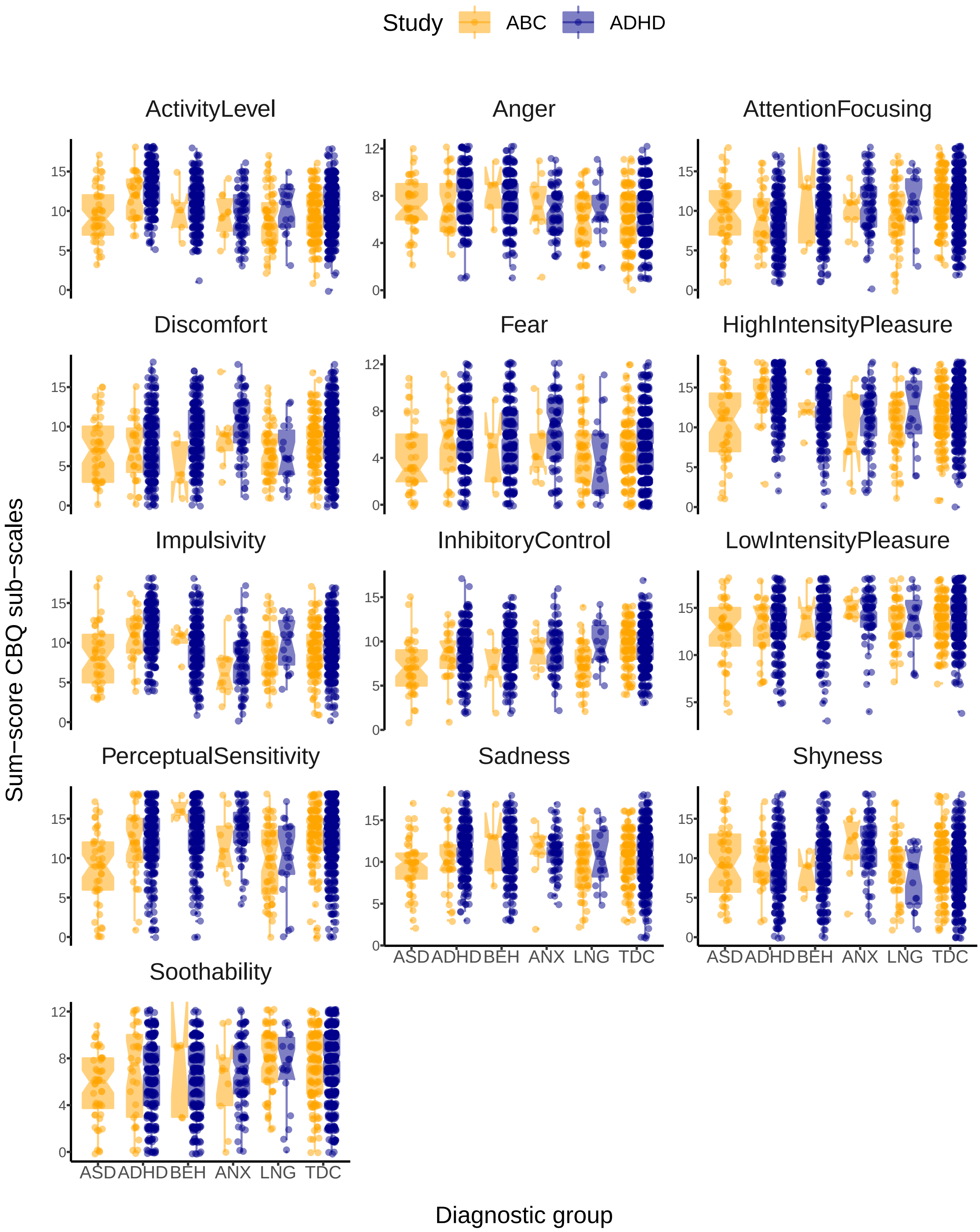


*Figure S17.* CBQ results in the ADHD and ABC studies, stratified by group.

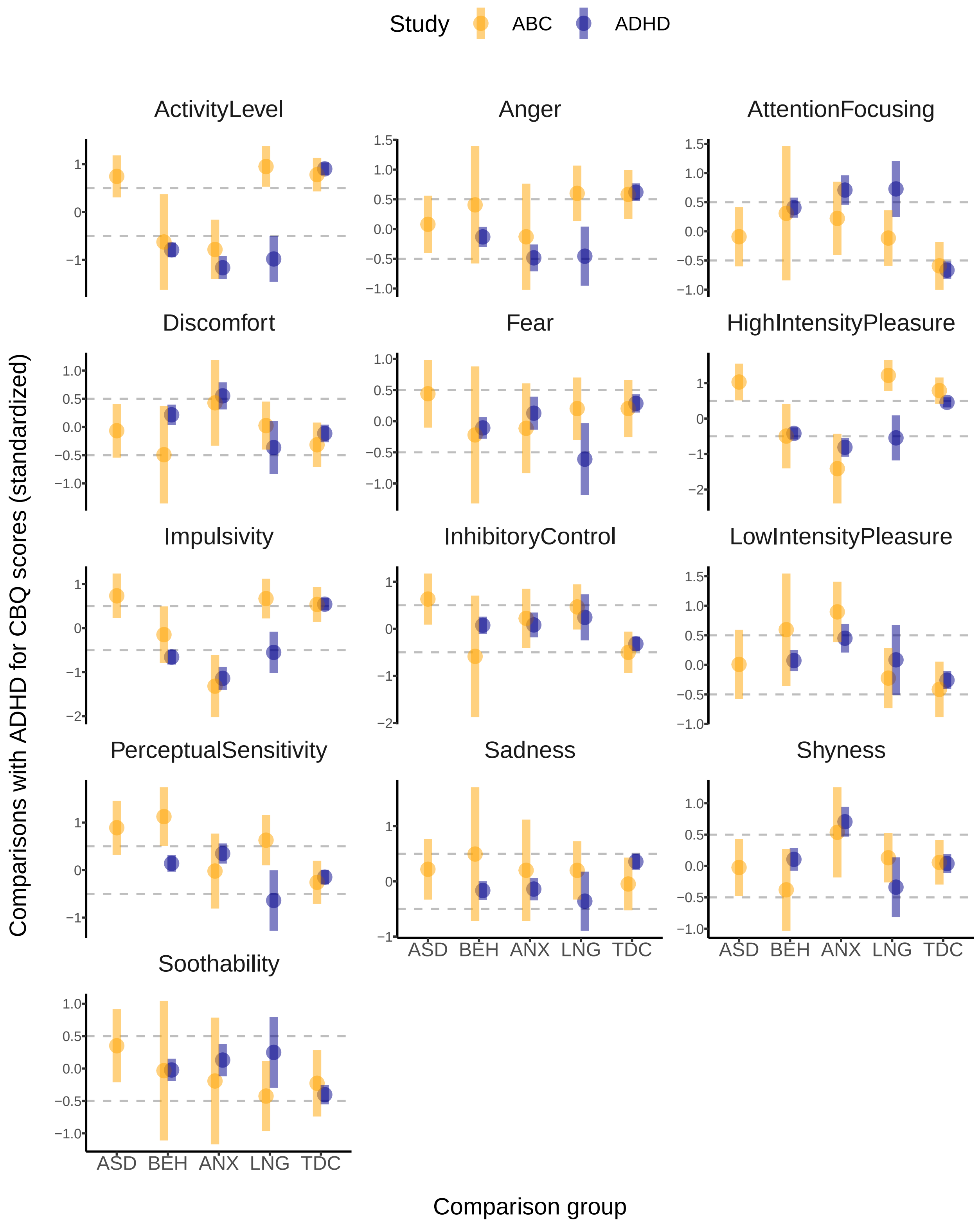


*Figure S18.* Comparison of children from ADHD and ABC Studies across CBQ results. All differences are based on scaled scores, whereby scaling was implemented by deviding the raw scores by the standardeviation of all scores (across sub-groups and studies).

Supplementary Material B

*
